# The impact of alcohol use in youth neurodevelopment: A systematic review of longitudinal structural neuroimaging studies

**DOI:** 10.64898/2026.01.06.26343559

**Authors:** Dhatsayini Rattambige, Govinda Poudel, Eugene McTavish, Ethan Murphy, Sunjeev K. Kamboj, Sarah Whittle, Valentina Lorenzetti

## Abstract

Youth alcohol use is a significant global health concern. Despite the widespread nature of alcohol use-related problems, the longitudinal effects of alcohol on brain structure in youth remain unclear. This review aimed to systematically synthesise the findings from the longitudinal structural magnetic resonance imaging (sMRI) literature on how alcohol use is associated with changes in brain structure in youth. Following PRISMA guidelines, five databases were searched, and studies of youth alcohol use that measured brain structure using sMRI at more than one time-point were included. A label-based meta-analysis (i.e., ratio of number of significant effects for a specific brain region to total number of analyses for that brain region) approach was employed to synthesise the findings. Sixteen studies were included. There was preliminary evidence that youth alcohol use is associated with reduced cortical volume (particularly in temporal regions) and attenuated increases in white matter volume over time. The role of pre-existing structural differences, and other moderating factors remains unclear due to limited research. Future longitudinal studies are needed to clarify the clinical significance of neurodevelopmental changes associated with youth alcohol use.

**Significance statement:** This systematic review synthesises evidence from 16 longitudinal neuroimaging studies on youth alcohol use and brain structure. Preliminary findings suggest adolescent drinking may be associated with reduced grey and white matter volume over time, with heavier consumption amplifying these effects. While methodological limitations prevent definitive conclusions, these potential neurodevelopmental disruptions during a critical brain maturation window could influence cognitive and behavioural outcomes. The review highlights the need for rigorous future research to further clarify the impact of alcohol on developmental brain trajectories, which could support targeted prevention approaches for adolescent brain health.

## 1. Introduction

Alcohol use among youth presents a significant public health challenge globally, with far-reaching consequences for individual well-being and societal health (WHO, 2024). The period of youth, defined as ages 12-24 (NIAA, 2023), is characterised by critical developmental changes in the brain and the formation of cognitive, emotional, and social skills (Giedd, 2008; Paus, 2005). During neurodevelopment, youth may be more susceptible to engaging in risky behaviours, such as binge drinking (WHO, 2024). Alcohol use by youth can also lead to serious consequences like driving under the influence and unintentional injuries (Hingson et al., 2003; Miller et al., 2007). The causes of youth alcohol use are multifaceted, rooted in a combination of biological, psychological, and social factors (Committee on Substance, 2010; Ryan et al., 2019). Meanwhile, the ongoing maturation of the brain during youth may increase susceptibility to the reinforcing effects of alcohol and may possibly increase risky alcohol use (Arain et al., 2013; Spear, 2015). Additionally, socio-environmental influences, such as peer pressure, family dynamics, and cultural attitudes towards alcohol, play a critical role in shaping drinking behaviours among youth (Jackson et al., 2014). Addressing these socio-environmental issues requires robust and extensive research to elucidate the association between alcohol use and neurodevelopment. Despite declines in some regions of the world, the prevalence of youth who drink alcohol remains high, especially in Eastern Europe and South America (Inchley et al., 2018). Therefore, understanding the impact of alcohol use on youth neurodevelopment is essential for informing prevention and intervention strategies that target young people.

Over the last decade, several theoretical models have been developed to explain the relationship between youth substance use and brain development (Baranger et al., 2023). One such model, the "dual-process" model, has been applied to understand risky behaviours in youth, including substance use (Casey et al., 2011; Steinberg, 2010). The model posits that striatal regions that support reward processing (e.g., nucleus accumbens, caudate, putamen) and other limbic areas implicated in emotion processing (e.g., amygdala, hippocampus) develop earlier than the prefrontal cortex, which underscores executive functions like planning and impulse control (Dudek et al., 2014; Evans & Stanovich, 2013). Therefore, according to the dual-process theory, the maturational discrepancy observed during neurodevelopment could lead to increased risk-taking and impulsive behaviour in youth (Shulman et al., 2016; Strang et al., 2013). Furthermore, animal and human research suggests that alcohol use during this developmental phase may be neurotoxic, which may further increase the risk of unhealthy alcohol use patterns (Lees et al., 2020; Spear, 2018; Squeglia & Gray, 2016).

Empirical studies in the last decade, using structural magnetic resonance imaging (sMRI), have provided consistent evidence of altered brain structure within frontal regions in youth alcohol users (Silveri et al., 2016). Interestingly, the frontal cortex is the last brain region to mature, and phenotypically, this can be associated with the delayed development of cognitive abilities, such as impulse control (Brumback et al., 2016; Casey et al., 2011). While extensive cross-sectional studies of alcohol consumption and youth brain structure have been conducted to date (Ewing et al., 2014; Spear, 2018), our understanding of how youth alcohol use is longitudinally associated with brain developmental trajectories remains limited. Importantly, longitudinal designs offer insights into the plausibility of directional and causal associations by elucidating the relationship between alcohol use and brain structure changes over time (Baranger et al., 2023; Nutt et al., 2021). Therefore, a better understanding of the longitudinal associations of alcohol use with neurodevelopment is extremely valuable for clinicians, researchers, and policy makers to create targeted interventions and policies (Squeglia & Gray, 2016). Although previous reviews have included longitudinal studies, they often lack a systematic and in-depth assessment of these studies and their nuances, such as variations in methodologies and the specific developmental stages examined (Cservenka & Brumback, 2017; Ewing et al., 2014; Hermens & Lagopoulos, 2018; Lees et al., 2018; Lees et al., 2019; Spear, 2018).

The primary aim of this review was to systematically synthesise the existing longitudinal sMRI research to identify the longitudinal associations between alcohol use and youth neuroanatomical development, including grey and white matter structure. The secondary aim was to synthesise research on how alcohol use metrics (e.g., dose, frequency, age of onset) and other moderating factors (e.g., sex, trauma, co-use of substances other than alcohol) at baseline and over time are associated with the reported neuroanatomical developmental trajectories.

## 2. Methods

### 2.1 Protocol and Registration

This review was pre-registered on the International Prospective Register of Systematic Reviews (submitted on 03/05/2023 and approved on 12/06/2023; ID: CRD42023418507). Each component of the review was performed in accordance with the latest Preferred Reporting Items for Systematic Reviews and Meta-Analyses (PRISMA) guidelines (Page et al., 2021).

### 2.2 Literature Search

A comprehensive systematic literature search was undertaken to identify studies using longitudinal sMRI to investigate the association between alcohol use and neuroanatomy. Articles published until January 6^th^ 2025, were included. Five databases were searched: PsychINFO (EBSCOhost), Medline (EBSCOhost), Scopus, PubMed and Embase. The search strategy consisted of four primary concepts: i) youth, ii) alcohol use, iii) neuroanatomy and iv) longitudinal study design. In each concept search, the keywords were combined with Medical Subject Headings (MeSH) using Boolean operators (e.g., OR/AND) and searched across study titles and abstracts. Selected studies were cross-referenced to ensure the inclusion of additional relevant work. The detailed search strategy is described in Supplementary Material. The final literature search was repeated prior to the finalisation of data analysis on August 5^th^, 2024

### 2.3 Study Selection

#### 2.3.1 Inclusion and exclusion criteria

The inclusion criteria were as follows: i) written in English language, ii) examined human participants, iii) peer-reviewed and empirical, iv) longitudinal study design, v) examined a group of participants who were alcohol users and a comparison group (i.e., healthy controls, non-binge drinkers, low users) as defined by each study’s protocol, vi) included participants up to the age of 24 years old at baseline; and vii) measured neuroanatomy using structural magnetic resonance imaging (sMRI). The exclusion criteria were: i) non-peer-reviewed, non-published (e.g., dissertations, conference abstracts, book chapters, or case reports), or non-empirical; ii) use of neuroimaging techniques other than sMRI; iii) included an adult sample of participants above the age of 24 at baseline; and iv) non-longitudinal studies (i.e., cross-sectional studies). The eligibility criteria included all studies published up to 2025, with no restrictions on the earliest publication year. Refer to Figure S1 for the PRISMA flowchart and for details on the search terms used.

### 2.4 Data extraction

Screening of articles using titles, abstracts, and full texts was completed in two stages. First, the titles and abstracts were screened by two independent raters (D.R., E.Mu.), who then resolved any discrepancies through discussion. Then, the full texts of all selected articles were screened by D.R. to ensure they met the inclusion criteria. During the latter process, articles were cross-referenced for additional relevant studies. One new study was identified via this process.

After applying the inclusion and exclusion criteria, 16 articles were eligible for data extraction. Data from the included 16 studies and 12 unique cohorts were systematically extracted and included: (i) sample characteristics; (ii) results for group differences; (iii) results for group (alcohol users vs non-users) by time interactions; (iv) results for dose-dependent alcohol use (e.g., high vs low alcohol use) by time interactions; and (v) results for additional included variables. Additional data that is not included in the main text includes: MRI magnet strength (tesla) and acquisition metrics (Table S1), and a sample of brain region categorisation approach as described in the data handling section (Table S2). Finally, results on group differences (i.e., cross-sectional comparisons between alcohol users and control groups) and additional moderating factors (i.e., sex, trauma, alcohol and cannabis co-use) are provided in Supplementary Section 2, as these represent secondary analyses beyond the primary focus on longitudinal brain developmental trajectories associated with alcohol use.

### 2.5 Risk of Bias and Quality Assessment

The risk of bias (low, moderate, high) of the reviewed literature was assessed using the National Institutes of Health, National Heart, Lung and Blood Institute: The Quality Assessment Tool for Observational Cohort and the Quality Assessment of Controlled Intervention Studies (National Heart, 2019). The risk of bias in the reviewed literature was determined in both assessment tools using fourteen distinct criteria, each of which was rated as “yes”, “no”, or “not applicable”. For each criterion, “yes” (i.e., no bias) equalled a score of 1 and “no” (i.e., bias present) a score of 0. The risk of bias was assessed for each study across all criteria. The studies were then classified using the mean bias score across all criteria, which was classified as either low (0.8-to-0.9 out of 1), moderately low (0.7), moderate (0.5-0.6) or high (<0.5) risk of bias.

### 2.6 Data synthesis

To examine the convergence of findings and conduct a quantitative synthesis, we employed a label-based meta-analysis approach (Laird et al., 2005; Phan et al., 2002). In the context of a neuroimaging review, a label-based approach refers to a simplified type of region of interest (ROI)- based meta-analysis, consisting of counting how many times a particular ROI is detected. This method was necessary due to the heterogeneity of the reviewed studies in terms of brain ROIs examined and the limitations in the information required to conduct a formal meta-analysis (Radua & Mataix-Cols, 2012).

We adapted the methodologies used in previous studies (Laird et al., 2005; Phan et al., 2002) to guide our label-based meta-analysis. That is, labels were derived directly from publications (author labels for ROIs). To summarise findings, these labels were combined into standard brain lobes (e.g., inferior frontal gyrus -> frontal, supramarginal gyrus -> parietal) (see Supplementary Table S2). Additionally, where studies investigated subcortical subregions, these were grouped by the overall subcortical region name (e.g., presubiculum -> hippocampus). The accuracy of this cataloguing process was verified by senior researchers (V.L., S.W., and E.Mc.).

An additional synthesis approach was adapted from a previous review (Rakesh & Whittle, 2021) where the consistency of findings was highlighted in the form of *m/n,* where *m* is the number of significant findings reported and *n* is the total number of instances of that ROI being investigated. To emphasise consistency, we defined a finding as a consistent finding if more than 50% of the studies reported a significant finding in a particular lobe (e.g., 3 out of 5 studies reported a significant finding in the frontal lobe). The findings are reported as percentages alongside their fractional equivalent throughout the results section, for example, “frontal lobe (60%, 3/5)”.

Moreover, results were reported for cohorts rather than by published study, treating each cohort as a separate finding. For example, if a study had three cohorts, each result was reported independently. Information on additional data handling is provided in the supplementary section. Finally, additional variables (e.g., co-use of substances other than alcohol, sex, trauma) were also extracted to explore the moderating effects of such variables, and the findings are qualitatively synthesised in the review.

## 3. Results

### 3.1 Study and participant characteristics

A total of 16 studies were included in this review (El Marroun et al., 2021; Hua et al., 2020; Infante et al., 2018; Infante et al., 2022; Jacobus et al., 2016; Jones et al., 2023; Luciana et al., 2013; Luo et al., 2022; Meda et al., 2017; Pérez-García et al., 2022; Pfefferbaum et al., 2018; Phillips et al., 2021; Squeglia et al., 2014; Squeglia et al., 2015; Sullivan et al., 2020; Sun et al., 2023) , which were based on data from 12 unique cohorts. Two of these studies (El Marroun et al., 2021; Jones et al., 2023) utilised data from multiple cohorts. Across the reviewed literature, seven studies sourced a subset of participants from a large parent database known as the National Consortium on Alcohol and Neurodevelopment in Adolescence (NCANDA) (Infante et al., 2022; Jones et al., 2023; Luo et al., 2022; Pfefferbaum et al., 2018; Phillips et al., 2021; Sun et al., 2023). In most cases, these studies were synthesised independently due to distinct neuroimaging outcomes investigated. However, if two or more studies using this cohort investigated the same neuroimaging outcome, it was counted only once. Furthermore, 14 out of 16 studies used data derived from the USA (i.e., participants were from the USA), and two studies used data from Europe (El Marroun et al., 2021; Pérez-García et al., 2022; Sullivan et al., 2020).

The demographic and alcohol use characteristics of the reviewed studies are summarised in Table 1. Studies included a total of 6,070 youth (females = 2,910; males = 3,160) aged between 9.9 – 20.0 years at baseline. Given that 7 studies (Infante et al., 2022; Jones et al., 2023; Luo et al., 2022; Pfefferbaum et al., 2018; Phillips et al., 2021; Sun et al., 2023) included participants from the NCANDA cohort and 2 studies (Squeglia et al., 2014; Squeglia et al., 2015) used the same cohort, the maximum number of unique participants included in this review is approximately 2,516. In terms of ethnicity, Caucasians were overrepresented, comprising about two-thirds of the participants, with four studies including only Caucasian participants (Infante et al., 2018; Jacobus et al., 2016; Squeglia et al., 2014; Squeglia et al., 2015).

**Table 1:**
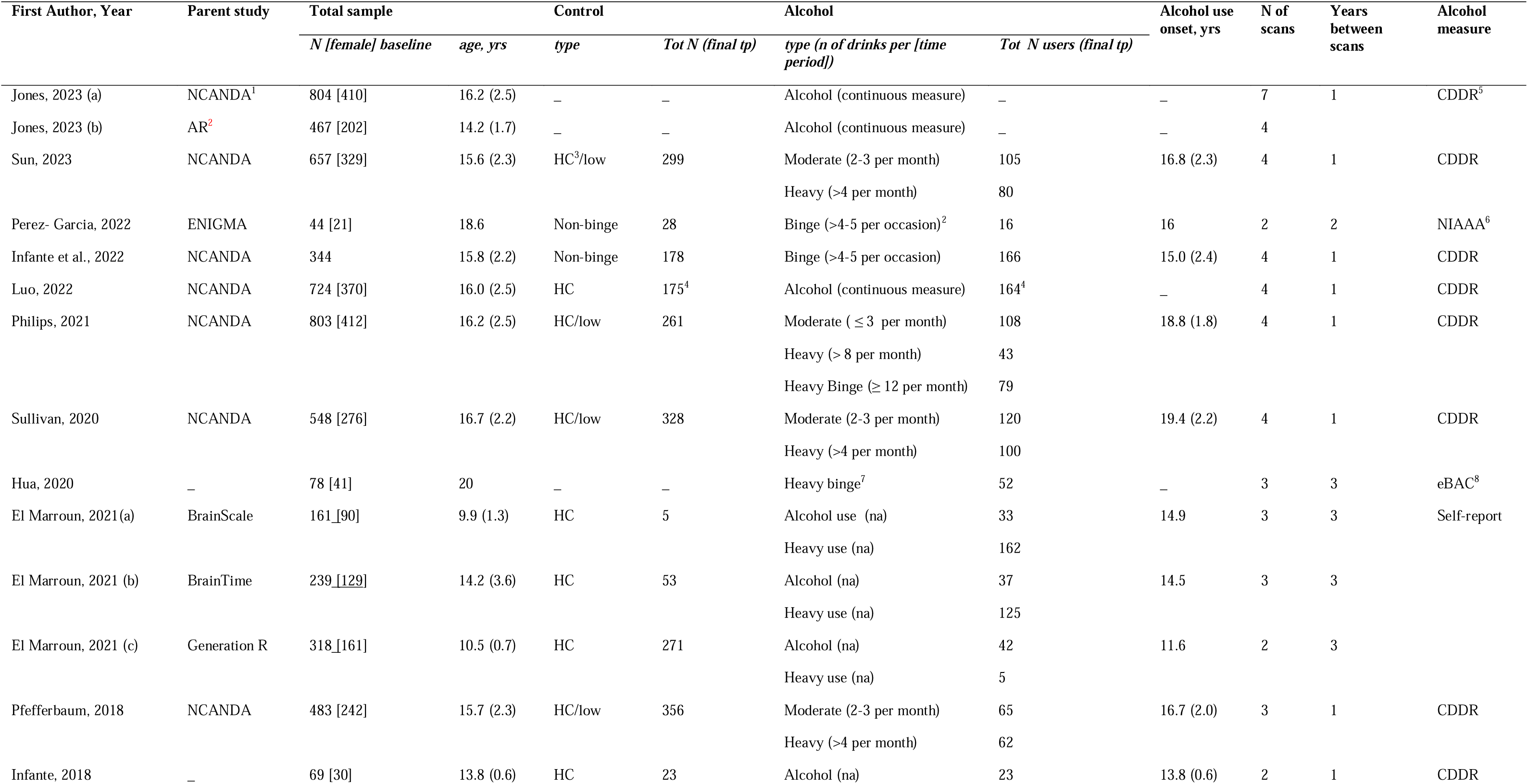
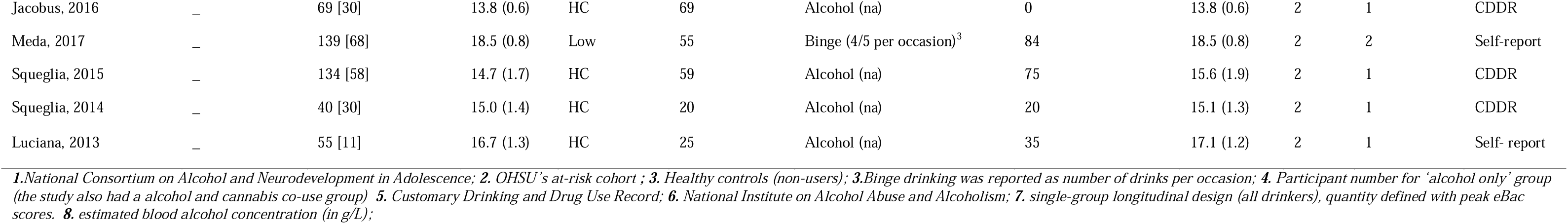
Overview of studies characteristics, sample demographics and alcohol use level across the reviewed literature.

| First Author, Year | Parent study | Total sample |  | Control |  | Alcohol |  | Alcohol use onset, yrs | N of scans | Years between scans | Alcohol measure |
| --- | --- | --- | --- | --- | --- | --- | --- | --- | --- | --- | --- |
|  |  | *N [female] baseline* | *age, yrs* | *type* | *Tot N (final tp)* | *type (n of drinks per [time period])* | *Tot N users (final tp)* |  |  |  |  |
| Jones, 2023 (a) | NCANDA<sup>1</sup> | 804 [410] | 16.2 (2.5) | – | – | Alcohol (continuous measure) | – | – | 7 | 1 | CDDR<sup>5</sup> |
| Jones, 2023 (b) | AR<sup>2</sup> | 467 [202] | 14.2 (1.7) | – | – | Alcohol (continuous measure) | – | – | 4 |  |  |
| Sun, 2023 | NCANDA | 657 [329] | 15.6 (2.3) | HC<sup>3</sup>/low | 299 | Moderate (2-3 per month) | 105 | 16.8 (2.3) | 4 | 1 | CDDR |
|  |  |  |  |  |  | Heavy (>4 per month) | 80 |  |  |  |  |
| Perez- Garcia, 2022 | ENIGMA | 44 [21] | 18.6 | Non-binge | 28 | Binge (>4-5 per occasion)<sup>2</sup> | 16 | 16 | 2 | 2 | NIAAA<sup>6</sup> |
| Infante et al., 2022 | NCANDA | 344 | 15.8 (2.2) | Non-binge | 178 | Binge (>4-5 per occasion) | 166 | 15.0 (2.4) | 4 | 1 | CDDR |
| Luo, 2022 | NCANDA | 724 [370] | 16.0 (2.5) | HC | 175<sup>4</sup> | Alcohol (continuous measure) | 164<sup>4</sup> | – | 4 | 1 | CDDR |
| Philips, 2021 | NCANDA | 803 [412] | 16.2 (2.5) | HC/low | 261 | Moderate ( ≤ 3 per month) | 108 | 18.8 (1.8) | 4 | 1 | CDDR |
|  |  |  |  |  |  | Heavy (> 8 per month) | 43 |  |  |  |  |
|  |  |  |  |  |  | Heavy Binge (≥ 12 per month) | 79 |  |  |  |  |
| Sullivan, 2020 | NCANDA | 548 [276] | 16.7 (2.2) | HC/low | 328 | Moderate (2-3 per month) | 120 | 19.4 (2.2) | 4 | 1 | CDDR |
|  |  |  |  |  |  | Heavy (>4 per month) | 100 |  |  |  |  |
| Hua, 2020 | – | 78 [41] | 20 | – | – | Heavy binge<sup>7</sup> | 52 | – | 3 | 3 | eBAC<sup>8</sup> |
| El Marroun, 2021(a) | BrainScale | 161 [90] | 9.9 (1.3) | HC | 5 | Alcohol use (na) | 33 | 14.9 | 3 | 3 | Self-report |
|  |  |  |  |  |  | Heavy use (na) | 162 |  |  |  |  |
| El Marroun, 2021 (b) | BrainTime | 239 [129] | 14.2 (3.6) | HC | 53 | Alcohol (na) | 37 | 14.5 | 3 | 3 |  |
|  |  |  |  |  |  | Heavy use (na) | 125 |  |  |  |  |
| El Marroun, 2021 (c) | Generation R | 318 [161] | 10.5 (0.7) | HC | 271 | Alcohol (na) | 42 | 11.6 | 2 | 3 |  |
|  |  |  |  |  |  | Heavy use (na) | 5 |  |  |  |  |
| Pfefferbaum, 2018 | NCANDA | 483 [242] | 15.7 (2.3) | HC/low | 356 | Moderate (2-3 per month) | 65 | 16.7 (2.0) | 3 | 1 | CDDR |
|  |  |  |  |  |  | Heavy (>4 per month) | 62 |  |  |  |  |
| Infante, 2018 | – | 69 [30] | 13.8 (0.6) | HC | 23 | Alcohol (na) | 23 | 13.8 (0.6) | 2 | 1 | CDDR |
| Jacobus, 2016 | — | 69 [30] | 13.8 (0.6) | HC | 69 | Alcohol (na) | 0 | 13.8 (0.6) | 2 | 1 | CDDR |
| Meda, 2017 | — | 139 [68] | 18.5 (0.8) | Low | 55 | Binge (4/5 per occasion)<sup>3</sup> | 84 | 18.5 (0.8) | 2 | 2 | Self-report |
| Squeglia, 2015 | — | 134 [58] | 14.7 (1.7) | HC | 59 | Alcohol (na) | 75 | 15.6 (1.9) | 2 | 1 | CDDR |
| Squeglia, 2014 | — | 40 [30] | 15.0 (1.4) | HC | 20 | Alcohol (na) | 20 | 15.1 (1.3) | 2 | 1 | CDDR |
| Luciana, 2013 | — | 55 [11] | 16.7 (1.3) | HC | 25 | Alcohol (na) | 35 | 17.1 (1.2) | 2 | 1 | Self-report |

*1. National Consortium on Alcohol and Neurodevelopment in Adolescence; 2. OHSU's at-risk cohort; 3. Healthy controls (non-users); 3. Binge drinking was reported as number of drinks per occasion; 4. Participant number for 'alcohol only' group (the study also had a alcohol and cannabis co-use group) 5. Customary Drinking and Drug Use Record; 6. National Institute on Alcohol Abuse and Alcoholism; 7. single-group longitudinal design (all drinkers), quantity defined with peak eBac scores. 8. estimated blood alcohol concentration (in g/L);*

**Table 2:** S**u**mmary **of results from group (i.e., alcohol use vs controls) by time interaction**

| Studies | Type of analysis | Groups compared | Neuroimaging metric | significant | non-significant |
| --- | --- | --- | --- | --- | --- |
| Perez-Garcia, 2022 | Whole brain and ROI | binge drinking vs non-binge | Surface area, GMV, thickness | – | Whole brain *and* ROI: **frontal** (middle, **cingulate** (anterior), **striatum** (accumbens)) |
| Luo, 2022* | ROI | Age (young vs old) & alcohol (heavy vs low) | GMV | *3-way interaction analysis between group, time and age - greater DECLINE over time in younger vs old drinkers in **temporal** (superior), & **parietal** (supramarginal)* | **frontal** (OFC, pars opercularis/orbitalis/triangularis pole, superior, caudal/rostral middle, pre/para/post-central), **hippocampus** (prosubiculum, subiculum proper, presubiculum and parasubiculum), **temporal** (pole, banks, entorhinal, middle/inferior, transverse), **cingulate** (caudal/rostral/posterior/anterior/ isthmus), **insula**, **occipital** (pericalcarine, fusiform, cuneus, lateral, lingual), **striatum** (accumbens/caudate/putamen), **parietal** (superior, precuneus) |
| Sullivan, 2020* | ROI | alcohol vs HC | GMV | *greater DECLINE over time in **cerebellum** lobes (I-IV, V, Vi, Crux I/II, VIIb) & vermis* | – |
|  |  |  | WMV | *smaller INCREASE over time in **cerebellum** vermis* | **cerebellum** lobes (I-IV, V, VI, Crux I/II, Viib) |
| El Marroun, 2021 (a) | Whole brain and ROI | alcohol vs HC | GMV | *greater DECLINE over time for total GMV and in **frontal** (lateral, medial, caudal middle, superior, rostral middle), **cingulate** (rostral anterior/posterior/isthmus), **amygdala**, **striatum** (caudate), **hippocampus*** | **striatum** (accumbens/putamen) |
|  |  |  | WMV | *smaller INCREASE in global white matter* | – |
| El Marroun, 2021 (b) |  | alcohol vs HC | GMV | *greater DECLINE over time for total GMV and in **frontal** (caudal middle), **cingulate** (rostral anterior/isthmus), **amygdala**, **hippocampus*** | **frontal** (lateral, medial, superior, rostral middle), **cingulate** (posterior), **striatum** (caudate) |
|  |  |  | WMV | – | **global white matter** |
| El Marroun, 2021(c) |  | alcohol vs HC | GMV | – | **total GMV** and **frontal** (lateral, medial, caudal middle, superior, rostral middle), **cingulate** (rostral anterior/posterior/isthmus), **amygdala**, **striatum** (accumbens/putamen/caudate), **hippocampus** |
|  |  |  | WMV | – | **global white matter** |

| Studies | Type of analysis | Groups compared | Neuroimaging metric | significant | non-significant |
| --- | --- | --- | --- | --- | --- |
| Meda, 2017 | Whole brain | binge drinking vs non-binge | GMV | *binge vs non-binge - greater DECLINE over time in **frontal** (paracentral pars opercularis/orbitalis/triangularis), **hippocampus** (parasubiculum), **cingulate** (anterior), **insula*** | n/a |
| Squeglia, 2015 | ROI | heavy drinking vs HC | GMV | *alcohol vs HC- greater DECLINE over time in **frontal** (lateral, medial, precentral), **hippocampus** (parasubiculum), **temporal** (superior, middle/inferior, transverse), **cingulate** (anterior), **insula*** | **frontal** (OFC/ pars opercularis/orbitalis/triangularis, pole, superior, caudal/rostral middle, pre/para/post-central), **occipital** (pericalcarine, fusiform, cuneus, lateral, lingual), **parietal** (supramarginal, superior, precuneus) |
|  |  |  | WMV | *heavy drinking vs HC - smaller INCREASE over time in **corpus callosum & pons*** | **central white matter** |
| Squeglia, 2014 | ROI | heavy drinking vs HC | GMV | *heavy drinking vs HC- greater DECLINE over time in **temporal** (inferior/middle), **striatum** (caudate), **brainstem*** | **frontal** (OFC/ pars opercularis/orbitalis/triangularis, pole, superior, caudal/rostral middle, pre/para/post-central), **cingulate** (caudal anterior/rostral anterior/posterior/anterior/isthmus) |
|  |  |  | WMV | *alcohol vs HC -smaller INCREASE over time in **cerebellum*** | **brainstem** |
| Luciana, 2013 | Whole brain | alcohol vs HC | thickness | *alcohol vs HC- greater DECLINE over time in **frontal** (Middle)* | n/a |
|  |  |  | WMV | *alcohol vs HC- smaller INCREASE over time in **frontal** (superior, precentral), **temporal** (middle), **occipital** (lingual), **cingulate gyrus*** | n/a |

**Abbreviations:** WMV = white matter volume ; GMV= grey matter volume; ‘\_’ = none, HC = healthy controls; OFC= orbitofrontal cortex (including : medial/lateral); IFG= inferior frontal gyrus (including: pars opercularis/orbitalis/triangularis); n/a = Not applicable as the analysis was whole brain voxel or vertex-wise ; ‘\*’ studies using NCANDA cohort

As illustrated in Figure 1, 12 of the 13 cohorts examined samples with an average age of 15.41 years at baseline (range 9.9 – 20) and an average age of 17.5 years by the second follow-up assessment across studies (range 13.0 – 21.0 years of age). Age of alcohol use onset was reported by 9 out of 13 cohorts (El Marroun et al., 2021(a&c); Hua et al., 2020; Infante et al., 2018; Infante et al., 2022; Jacobus et al., 2016; Luciana et al., 2013; Meda et al., 2017; Pérez-García et al., 2022; Pfefferbaum et al., 2018; Phillips et al., 2021; Squeglia et al., 2014; Squeglia et al., 2015; Sullivan et al., 2020; Sun et al., 2023). The average age of alcohol use onset across the reviewed studies was 15.8 years (range 11.6 to 19.4). Only four studies had alcohol naïve participants at baseline to interpret the effects of youth alcohol use initiation on structural brain development (Hua et al., 2020; Infante et al., 2022; Meda et al., 2017; Pérez-García et al., 2022)

**Figure 1.**
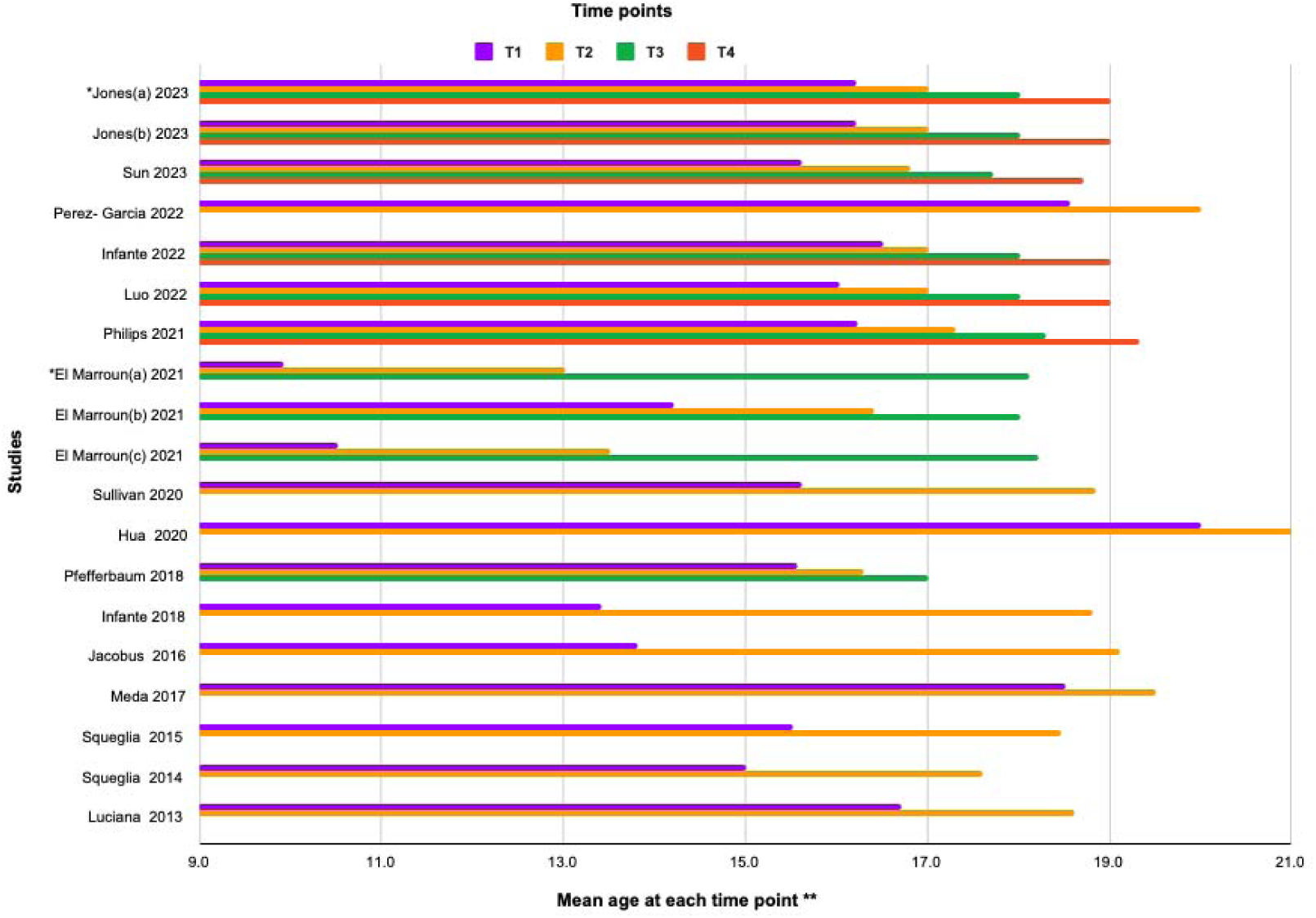
Mean age at each time point across longitudinal studies *Note.* The graph shows the number of time points as reported in each study. It also shows how age progresses across these time points as reported in each cohort included within a study. *Jones et al. (2023 and El Marroun et al. (2021 had reported findings within different cohorts; the various cohorts are distinguished alphabetically (a-c). *\*\**The mean age can be determined at the point where the bar ends.

### 3.2 Interaction of group (alcohol use vs controls) by time

Group (alcohol use vs control) by time interaction effects were reported for nine unique cohorts (El Marroun et al., 2021(a-c); Luciana et al., 2013; Luo et al., 2022; Meda et al., 2017; Pérez-García et al., 2022; Squeglia et al., 2014; Squeglia et al., 2015; Sullivan et al., 2020), details of which can be seen in Table 2. The most studied imaging metric was GMV (n=9), followed by WMV (n=7), cortical thickness (n=2) and surface area (n=1). For a visual summary of the most consistent group-by-time interaction effects, refer to Figure 2.

**Figure 2.**
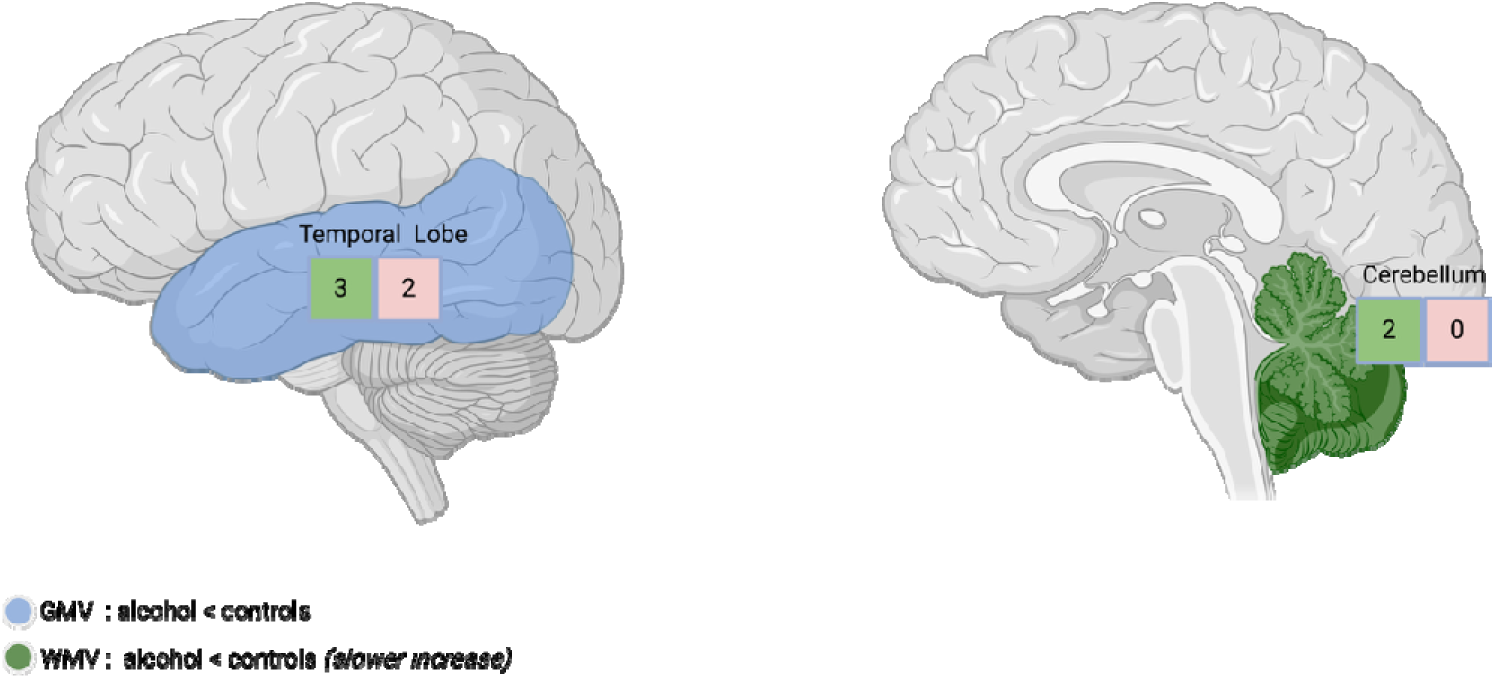
Significant group-by-time interaction effects for alcohol use versus controls *Note.* This figure summarises the most consistent findings from studies investigating group (alcohol use vs. controls) by time interaction effects on brain structure, highlighting both grey matter volume (GMV) and white matter volume (WMV) results. The most consistent significant findings were reported in the temporal lobe and cerebellum. The colour of the boxes indicates statistical significance: green boxes represent significant results, and pink boxes represent non-significant results. The numbers inside each box reflect the number of studies that found significant versus non-significant group-by-time interactions in each brain region.

***GMV***: The most consistent finding was a significant GMV decline over time in alcohol-using groups (e.g., heavy and binge drinkers) compared to controls (e.g., healthy controls/non-users, non-binge drinkers), which was reported in 8 independent cohorts. The group-by-time effects were most commonly evident in regions within the temporal lobe (60%, 3/5). One study reported that the effect of alcohol use on GMV decrease was attenuated among older alcohol-using participants compared to the younger participants, within the temporal (superior) and parietal (supramarginal) regions (Luo et al., 2022).

***WMV:*** In seven cohorts, the increase of WMV over time was less marked in the alcohol than in the control groups. This effect was most consistently located within the cerebellum (100%, 2/2) (Squeglia et al., 2014; Sullivan et al., 2020).

***Cortical thickness:*** Only two cohorts examined group-by-time effects on cortical thickness. The results were mixed, with the middle frontal gyrus being affected in one study (Pérez-García et al., 2022) but not another study Luciana et al. (2013).

***Surface area:*** A single study examined group-by-time effects on surface area within the frontal and cingulate cortices, whereby groups were binge drinkers and non-binge drinkers. The study reported non-significant effects (Pérez-García et al., 2022).

### 3.3 Alcohol dose-dependent longitudinal effects

This section and Table 3 synthesise the results from the ten cohorts for which the interaction between alcohol ‘dose’ (e.g., low, moderate, heavy, binge) and time were investigated (El Marroun et al., 2021(a-c); Infante et al., 2018; Meda et al., 2017; Pérez-García et al., 2022; Pfefferbaum et al., 2018; Phillips et al., 2021; Sullivan et al., 2020; Sun et al., 2023). Alcohol consumption level, measured by duration, dose, and frequency, was classified into subgroups in some studies (e.g., low, moderate, heavy, and binge drinkers; intoxicated vs. non-intoxicated). In other studies, alcohol use level was measured on a continuous scale (e.g., number of drinks consumed). The most consistently studied metric of brain structure was GMV (n=8), followed by WMV (n= 5), cortical thickness (n=1) and surface area (n=1). For a visual summary of the most consistent finding of the longitudinal dose-dependent effects, refer to Figure 3.

**Figure 3.**
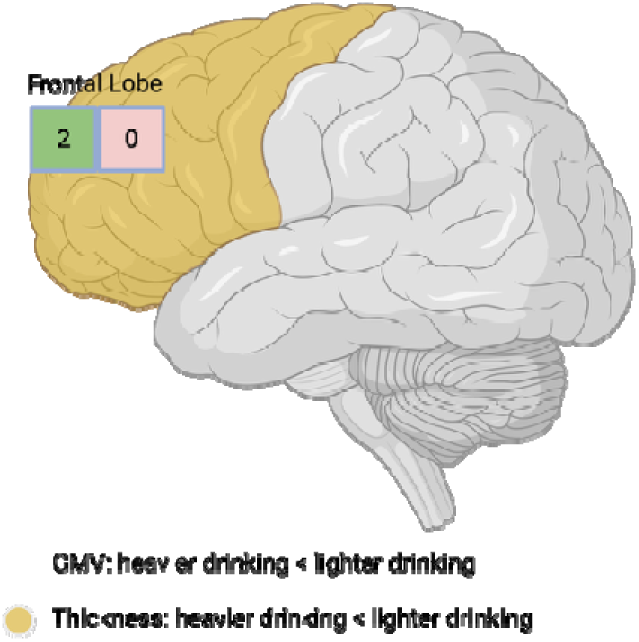
Brain regions showing dose-dependent longitudinal effects of alcohol use *Note.* This figure summarises the most consistent findings reported in studies investigating the dose-dependent longitudinal effects of alcohol on brain structure, highlighting cortical thickness results. The colour of the boxes indicates significance: green for significant results and pink for non-significant results. The numbers inside each box reflect the number of studies that found significant and non-significant effects.

**Table 3.**
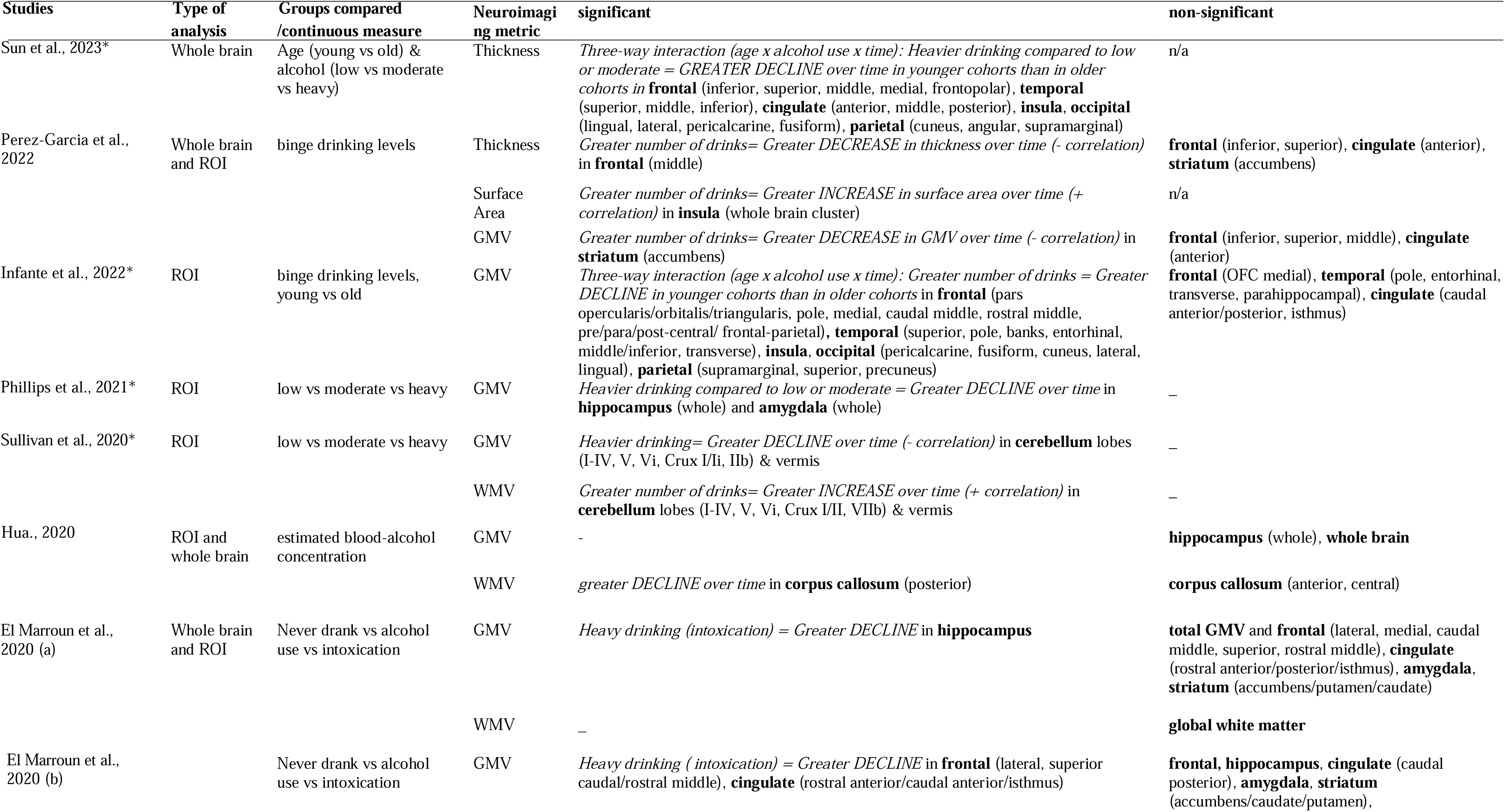
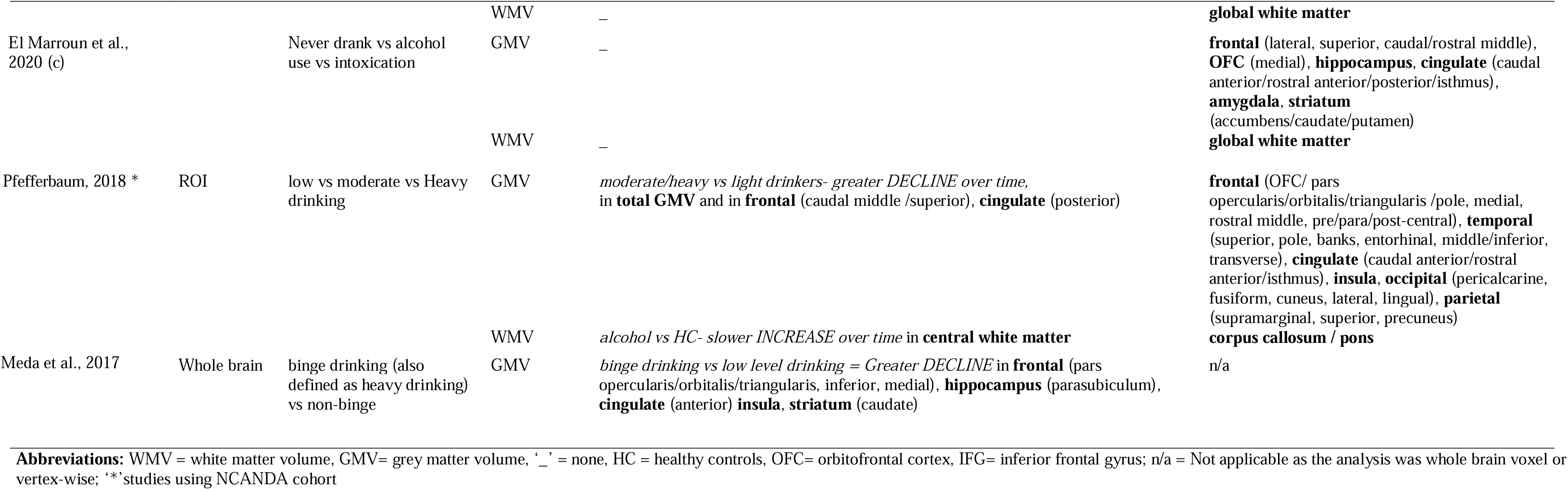
Overview of findings on dose-dependent longitudinal effects

***GMV:*** In seven out of 8 cohorts, heavier drinking was associated with a greater decline of GMV over time (El Marroun et al., 2021(a-c); Infante et al., 2022; Meda et al., 2017; Pérez-García et al., 2022; Pfefferbaum et al., 2018; Phillips et al., 2021; Sullivan et al., 2020). While there were no brain regions/lobes consistently implicated, the decline in GMV was reported within the frontal lobe in 3/6 cohorts. Of note, one study (Infante et al., 2022) reported a significant interaction between binge drinking episodes in the past year, time, and baseline age. The results indicated that GMV declined over time in various regions (e.g. frontal, temporal and cingulate) as a function of more binge drinking episodes, particularly in younger individuals.

***WMV:*** There was largely non-significant evidence that alcohol use levels were associated with WMV changes over time (El Marroun et al., 2021(a-c); Pfefferbaum et al., 2018; Sullivan et al., 2020).

***Cortical thickness:*** There was emerging yet consistent association between greater number of alcohol consumption and greater decline in frontal cortical thickness (100%, 2/2), especially within the frontal middle gyrus. There were mixed results for the cingulate cortex: Sun et al. (2023) observed significant declines in heavy alcohol users compared to moderate users, whereas Pérez-García et al. (2022) found no significant effects between different levels of binge drinkers.

***Surface area:*** One study reported that greater occasions of binge drinking positively correlated with increased surface area in the insula over time (Pérez-García et al., 2022).

### 3.4 Baseline differences between alcohol users and healthy controls

As shown in Table 4, four studies reported on baseline differences (Jacobus et al., 2016; Meda et al., 2017; Pfefferbaum et al., 2018; Squeglia et al., 2014). Two of the four studies did not examine alcohol naïve participants at baseline as their control groups were at least low-level drinkers at baseline (Meda et al., 2017; Pfefferbaum et al., 2018). GMV was the most consistently measured brain metric (n=3), followed by WMV (n=2) and cortical thickness (n=1). The most consistently reported significant findings were observed within the frontal lobe (66%, 2/3) and the cingulate (66%, 2/3), where smaller GMV was reported in alcohol use initiators (Squeglia et al., 2014) and binge drinkers (Meda et al., 2017) compared to non-users or non-binge drinkers. Lastly, Jacobus et al. (2016) reported greater thickness at baseline in alcohol users only compared to the other groups (i.e., alcohol + cannabis and controls).

**Table 4:** B**a**seline **difference between alcohol users and non-users**

| Studies | Analysis used | Groups compared | Neurometric | significant | non-significant |
| --- | --- | --- | --- | --- | --- |
| Pfefferbaum et al., 2018 | ROI | low vs moderate vs heavy drinking (measured after alcohol use onset) | GMV | – | **frontal** (OFC/ pars opercularis/orbitalis/triangularis /pole, medial, caudal middle, rostral middle, pre/para/post-central), **temporal** (superior, pole, banks, entorhinal, middle/inferior, transverse), **cingulate** (caudal anterior/posterior, rostral anterior/isthmus), **insula**, **occipital** (pericalcarine, fusiform, cuneus, lateral, lingual), **parietal** (supramarginal, superior, precuneus) |
|  |  |  | WMV | – | **global white matter, pons, corpus callosum** |
| Jacobus et al., 2016 | ROI | alcohol vs HC vs co-users | thickness | *GREATER in alcohol initiators vs HC and co-users in **frontal** (pars triangularis, superior, pre/para central, rostral middle), **hippocampus** (parasubiculum), **parietal** (inferior, supramarginal, superior), **insula*** | **frontal** (pole), **occipital** (pericalcarine, fusiform, cuneus, lateral, lingual), **parietal** (precuneus) |
| Meda et al., 2017 | Whole brain | binge vs non-binge drinking (measured after alcohol use onset) | GMV | *SMALLER in binge drinkers vs non -binge in **frontal** (superior, inferior, middle), **hippocampus** (parasubiculum), **cingulate** (anterior), **insula*** | n/a |
| Squeglia et al., 2014 | ROI | initiators vs HC | GMV | *SMALLER in heavy drinkers vs HC in **cingulate** (rostral anterior / caudal anterior, isthmus), **frontal** (pars triangularis)* | **frontal** (OFC/ pars opercularis/orbitalis, pole, superior, caudal/rostral middle, pre/para/post-central), **temporal** (inferior/middle), **striatum** (caudate), **brainstem, cingulate** (posterior/anterior/isthmus) |
|  |  |  | WMV | *SMALLER in alcohol initiators vs HC in **cerebellum*** | **brainstem** |

**Abbreviations:** WMV = white matter volume, GMV= grey matter volume, ‘–’ = none, HC = healthy controls, OFC= orbitofrontal cortex, IFG= inferior frontal gyrus; n/a = Not applicable as the analysis was whole brain voxel or vertex-wise

### 3.5 Additional Moderators

Eight studies investigated additional variables as moderators of the longitudinal relationship between alcohol use and structural neurodevelopment in youth (Infante et al., 2018; Infante et al., 2022; Jacobus et al., 2016; Jones et al., 2023; Luo et al., 2022; Pfefferbaum et al., 2018; Sullivan et al., 2020; Sun et al., 2023). These variables include family history of alcohol use or alcohol use disorder (AUD) (n=3), co-use of alcohol and cannabis (n=3), sex (n=2), and trauma (n=2). Findings for the former two moderators are described below. For a comprehensive examination of moderators tested in fewer than three studies (i.e., sex and trauma), refer to Supplementary Section 2.

Family history of AUD was a consistent moderator investigated across the NCANDA studies (Pfefferbaum et al., 2018; Sullivan et al., 2020; Sun et al., 2023). Two studies showed that alcohol consumption exacerbated GMV decline in youth with a positive family history of AUD (Pfefferbaum et al., 2018; Sullivan et al., 2020). The third study (Sun et al., 2023) found no moderating role of family history of AUD on cortical thickness changes associated with youth alcohol use. Alcohol and cannabis co-use was also examined across three studies. Infante et al. (2018) and Jacobus et al. (2016) found that alcohol-only users showed greater reductions in the surface area and cortical thickness of the frontal and parietal lobes over time compared to co-users. Conversely, Luo et al. (2022) reported that co-users experienced significantly greater declines in brain volume in the frontal, temporal, and parietal lobes compared to both non-users and alcohol-only users.

### 3.6 Risk of bias

The reviewed literature demonstrated varying levels of risk of bias, categorised into low, moderately low, and moderate risk based on specific criteria as demonstrated in Table 5. The average within-cohort risk of bias was 0.7, and the average between-cohort risk of bias was 0.5.

**Table 5:** Risk of bias using the National Institutes of Health, National Heart, Lung and Blood Institute

|  | Jones 2023 (a) | Jones 2023 (b) | Sun 2023 | Perez-Garcia 2022 | Infante 2022 | Luo 2022 | Phillips 2021 | Sullivan 2019 | Hua 2021 | El Marroun 2021 (a) | El Marroun 2021 (b) | El Marroun 2021 (c) | Pfefferbaum 2018 | Infante 2018 | Jacobus 2016 | Meda 2017 | Squeglia 2015 | Squeglia 2014 | Luciana 2013 | Average between-study item score |
| --- | --- | --- | --- | --- | --- | --- | --- | --- | --- | --- | --- | --- | --- | --- | --- | --- | --- | --- | --- | --- |
| **A** | 1 | 1 | 1 | 1 | 1 | 1 | 1 | 1 | 1 | 1 | 1 | 1 | 1 | 1 | 1 | 1 | 1 | 1 | 1 | 1.0 |
| **B** | 1 | 0 | 1 | 1 | 1 | 1 | 1 | 1 | 1 | 1 | 1 | 1 | 1 | 1 | 1 | 1 | 1 | 1 | 1 | 0.9 |
| **C** | 0 | 0 | 1 | 1 | 1 | 1 | 1 | 1 | 1 | 1 | 1 | 1 | 1 | 1 | 1 | 1 | 1 | 1 | 1 | 0.9 |
| **D** | 1 | 1 | 1 | 1 | 1 | 1 | 1 | 1 | 1 | 1 | 1 | 1 | 1 | 1 | 1 | 1 | 1 | 1 | 1 | 1.0 |
| **E** | 1 | 1 | 1 | 1 | 1 | 1 | 1 | 1 | 1 | 1 | 1 | 1 | 1 | 1 | 1 | 1 | 1 | 1 | 1 | 1.0 |
| **F** | 0 | 0 | 0 | 0 | 0 | 0 | 0 | 0 | 0 | 0 | 0 | 0 | 0 | 0 | 0 | 0 | 0 | 0 | 0 | 0.0 |
| **G** | 1 | 1 | 1 | 1 | 1 | 1 | 1 | 1 | 1 | 1 | 1 | 1 | 1 | 1 | 1 | 1 | 1 | 1 | 1 | 1.0 |
| **H** | 1 | 0 | 1 | 1 | 1 | 1 | 1 | 1 | 0 | 1 | 1 | 1 | 1 | 1 | 1 | 1 | 1 | 1 | 1 | 0.9 |
| **I** | 1 | 1 | 1 | 1 | 1 | 1 | 1 | 1 | 0 | 0 | 0 | 0 | 1 | 0 | 0 | 1 | 0 | 0 | 0 | 0.5 |
| **J** | 0 | 0 | 0 | 0 | 0 | 0 | 0 | 0 | 0 | 0 | 0 | 0 | 0 | 0 | 0 | 0 | 0 | 0 | 0 | 0.0 |
| **K** | 1 | 0 | 1 | 1 | 1 | 1 | 1 | 1 | 0 | 1 | 1 | 1 | 1 | 1 | 1 | 1 | 1 | 1 | 1 | 0.9 |
| **L** | 1 | 1 | 1 | 1 | 1 | 1 | 1 | 1 | 1 | 1 | 1 | 1 | 1 | 1 | 1 | 1 | 1 | 1 | 1 | 1.0 |
| **M** | 0 | 0 | 0 | 0 | 0 | 0 | 0 | 0 | 0 | 0 | 0 | 0 | 0 | 0 | 0 | 0 | 0 | 0 | 0 | 0.0 |
| **N** | 0 | 0 | 0 | 1 | 1 | 1 | 0 | 1 | 1 | 1 | 1 | 1 | 0 | 0 | 0 | 0 | 0 | 0 | 0 | 0.4 |
| **O** | 1 | 1 | 1 | 1 | 1 | 1 | 1 | 1 | 0 | 1 | 1 | 0 | 1 | 1 | 1 | 1 | 1 | 1 | 1 | 0.9 |
| Within-study risk of bias | 0.7 | 0.5 | 0.7 | 0.8 | 0.8 | 0.8 | 0.7 | 0.8 | 0.5 | 0.7 | 0.7 | 0.7 | 0.7 | 0.7 | 0.7 | 0.7 | 0.7 | 0.7 | 0.7 |  |

**Notes of full questions:** **A.** Research question/objective clearly stated ; **B.** Study population clearly specified and defined; **C.** Participation rate was at least 50% **D.** Subjects are selected or recruited from similar populations; **E.** Inclusion and exclusion criteria pre-specified and applied uniformly ; **F.** Sample size justified and power description provided; **G.** Exposure is measured prior to the outcome; **H.** Timeframe is sufficient to expect an association; **I.** Different level of exposures are examined; **J.** Exposure measures are clearly defined and reliable; **K.** Exposure is measured more than once over time; **L.** Outcome measures are clearly defined and reliable; **M.** outcome assessors are blinded to exposure status; **N.** Loss to follow-up after baseline is 20% or less; **O.** Key confounders are measured and statistically adjusted

All 16 studies clearly stated their research questions and objectives, specified and defined study populations, applied inclusion and exclusion criteria uniformly, measured exposures before outcomes and clearly defined outcome measures. Finally, one study had a follow-up period of five weeks between neuroimaging sessions, which may not fully capture the effects on neurodevelopment (Squeglia et al., 2009). However, none of the studies provided sample size justifications or power analyses, nor did they ensure that exposure measures were consistently defined or that outcome assessors were blinded. Additionally, loss to follow-up was a significant issue, with only some studies maintaining less than 20% loss. The most significant biases across the studies were the lack of justification for sample size, inadequate descriptions of power, and unreliable exposure measures, which contributed to heterogeneity in how alcohol use was measured. Figure 4 illustrates the different methods used to measure alcohol use across studies, highlighting variations in frequency and dose measurements in each study.

**Figure 4.**
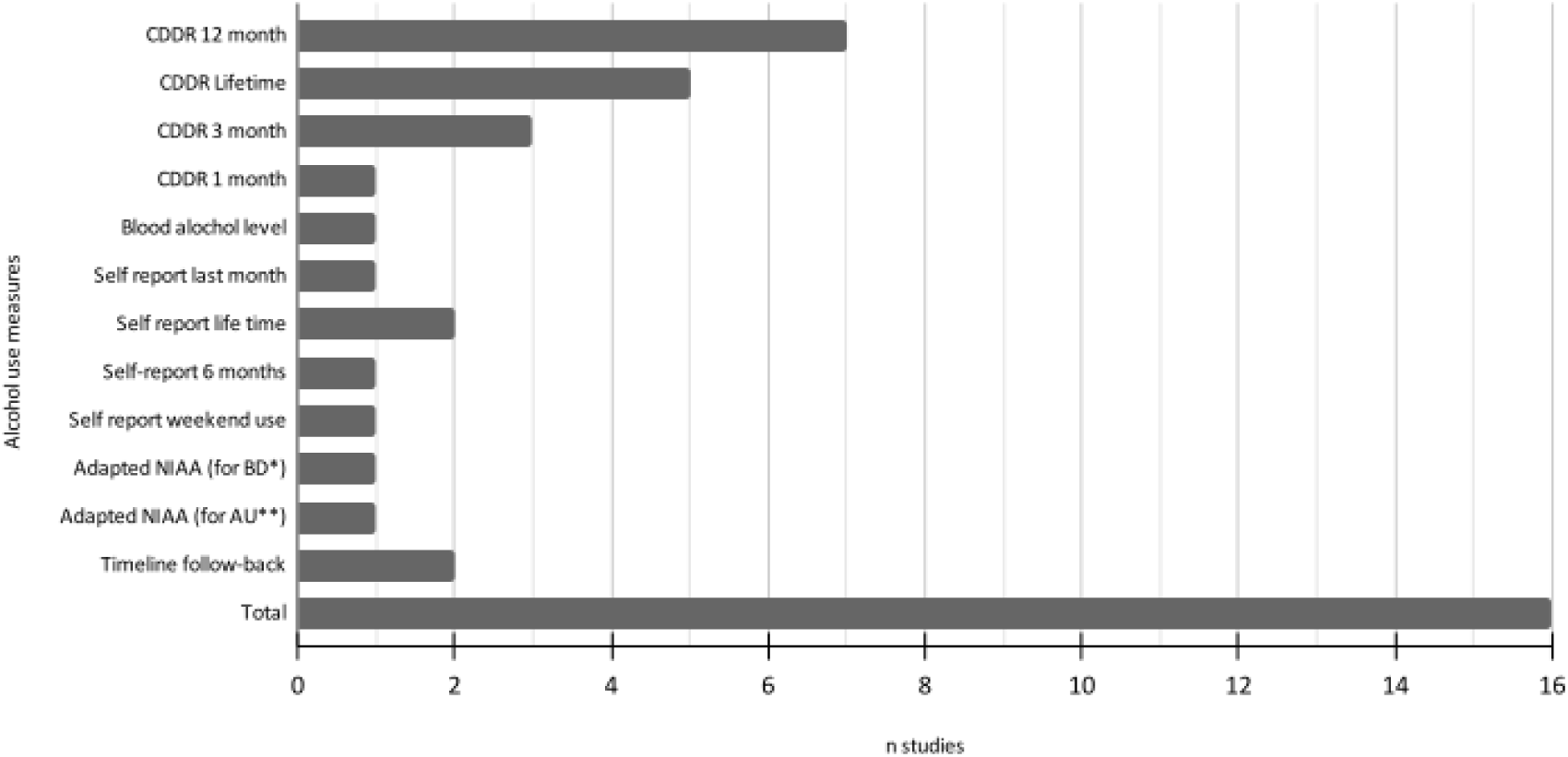
Alcohol use measurement methods across the literature *Note.* CDDR = Customary Drinking and Drug Use Record; NIAA = National Institute on Alcohol Abuse and Alcoholism; BD* = Binge Drinking; AU** = Alcohol Use. Multiple studies used more than one measurement method, contributing to the heterogeneity in exposure assessment identified as a significant source of bias.

## 4. Discussion

Overall, across the 13 cohorts, the most consistently reported finding was an interaction effect between alcohol use (i.e., alcohol users versus controls) and time, whereby alcohol use was associated with reductions in GMV and attenuation of WMV increases over time, with the GMV effect most consistently reported in temporal regions. A dose-dependent longitudinal effect of alcohol on brain structure was also apparent, although there was no specific brain region that was consistently implicated. Few studies investigated differences in brain structure prior to alcohol use initiation, or factors that may moderate the effect of alcohol use on brain structure over time. While further research is needed, there was emerging preliminary evidence for pre-existing structural differences, and moderating effects of age, cannabis co-use and family history.

### 4.2 Longitudinal effects of alcohol use on brain structure

The reviewed longitudinal studies suggested that alcohol consumption is associated with a significant decline in GMV over time. This effect was particularly consistent within the temporal lobe; however, this was only evidenced in three cohorts, hence it cannot be considered conclusive. Evidence for age-related differences was also limited, with one study (Infante et al., 2022) showing a more pronounced GMV decline in younger binge drinkers. Attenuated WMV increases over time in alcohol users were observed, with these effects observed primarily in the cerebellum; however, with few studies investigating the cerebellum, the findings are limited and should be interpreted cautiously.

The temporal lobe is crucial for functions such as memory processing, language comprehension, and emotional regulation, which continue to develop during adolescence (Gogtay et al., 2004). Systematic reviews of cross-sectional studies consistently associate alcohol use with reduced GMV in these regions (Carbia et al., 2018; Lees et al., 2020; Lees et al., 2019). The longitudinal findings build on this understanding, suggesting that GMV reductions in alcohol users may reflect deviations from typical brain development during youth (Gogtay et al., 2004; Volkow et al., 2014). However, the temporal dynamics of these structural changes are not yet fully understood. Further, it remains uncertain whether these alterations are directly linked to cognitive changes, whether they are transient or enduring, and to what extent they can be attributed specifically to adolescent alcohol use. Additionally, the impact of varying levels of alcohol consumption on these effects has yet to be determined.

### 4.2 Dose-dependent longitudinal effects of alcohol on brain structure

There was consistent evidence that higher ‘doses’ of alcohol were associated with greater brain GMV decline. Preliminary data suggest that heavier alcohol use during adolescence may be associated with more pronounced GMV decline in the frontal lobe, a region critical for executive functions and decision-making. However, these findings require further validation. The observed greater GMV decreases in frontal regions may have implications for cognitive (e.g., cognitive flexibility, inhibitory control), emotional (e.g., emotional regulation), and behavioural outcomes (e.g., academic performance) (Cservenka & Brumback, 2017; Snyder et al., 2017; Suñol et al., 2022). However, the current evidence base is constrained by the limited number of independent cohorts. Additional research is necessary to establish the relationship between structural alterations and cognitive (and other) outcomes (Cservenka & Brumback, 2017; Hingson et al., 2003; Lees et al., 2020; Mota et al., 2013).

### 4.3 Effects of alcohol use between younger and older youth

Two studies observed age-related differences in the impact of alcohol use over time, highlighting a possible heightened susceptibility of younger individuals to the effects of alcohol (Infante et al., 2022; Luo et al., 2022). While attenuated effects in older youth might suggest compensatory mechanisms, the interpretation of age-specific vulnerability requires careful consideration (Foulkes & Blakemore, 2018; Giedd, 2008). Although younger individuals demonstrate higher neuroplasticity during neurodevelopment (Fuhrmann et al., 2015; Sullivan & Pfefferbaum, 2019), brain development follows individualised trajectories influenced by multiple biological and environmental factors. This developmental plasticity may also facilitate recovery; emerging evidence suggests cognitive improvements following alcohol abstinence during development (Lees et al., 2020; Squeglia et al., 2017), although the corresponding structural recovery remains unexplored. Finally, although age-related susceptibility to the effects of alcohol on neurodevelopment may be clinically significant, it should not oversimplify developmental vulnerability, as neural sensitivity varies across brain systems (Andersen, 2003) and individual developmental trajectories (Fuhrmann et al., 2015; Spear, 2015).

### 4.4 Baseline differences predating youth alcohol use

Among the four studies examining baseline structural differences between alcohol users and controls, two studies included alcohol-naïve participants at baseline (Jacobus et al., 2016; Squeglia et al., 2014). Jacobus et al. (2016) reported greater thickness at baseline in alcohol users compared to other groups, such as co-users and controls. Whereas, Squeglia et al. (2014) a smaller GMV was reported in alcohol use initiators compared to non-users. These results suggest potential structural brain differences that may exist before alcohol use begins. However, several limitations affect the interpretation of these findings. For example, the studies used different metrics, such as cortical thickness and GMV, making direct comparison difficult. Existing cross-sectional literature has identified significant correlations between brain morphology in specific fronto-striatal regions and the cerebellum with future alcohol use behaviours (Kühn et al., 2019; Rane et al., 2022; Seo et al., 2019; Whelan et al., 2014), suggesting that brain morphology could potentially serve as a biomarker for later alcohol use. However, these correlations do not necessarily imply predictive capability.

### 4.5 Additional moderators influencing the association between alcohol use and neurodevelopment

The role of the additional moderators was reviewed to further elucidate the longitudinal relationship between youth alcohol use and structural brain development. Emerging findings suggest that alcohol use could be associated with greater reductions in brain volume, surface area, and thickness compared to cannabis co-users and non-users. Whilst not conclusive, these findings could indicate that alcohol, when used alone, has a more substantial impact on brain structure than when used in addition to cannabis. To truly establish the mechanism of the relationship between alcohol and cannabis co-use, it is essential to understand the specific quantities of cannabis and alcohol consumed together. The effects of co-use may involve complex interactions between these substances, which have been shown to produce distinct neural effects compared to single substance use (Karoly et al., 2015; Lisdahl et al., 2013), yet these are not yet fully understood from a neuropharmacological perspective (Bedillion et al., 2021; Infante et al., 2020). More studies, especially longitudinal investigations tracking the temporal progression of these interactions (Yurasek et al., 2017), are needed to confirm these effects and to understand the precise mechanisms behind the observed interactions.

Two out of three studies investigating familial history of AUD reported significant moderating effects (Pfefferbaum et al., 2018; Sullivan et al., 2020). While these studies investigated the same sample, these emerging findings align with existing literature indicating that youth with a family history of AUD may be more vulnerable to alcohol-related effects on brain structure and cognition (Elliott et al., 2012; Haeny et al., 2020; Walters, 2002). This effect may be driven by genetic predispositions and/or disruptions in the caregiving environment, which influence neurobiological mechanisms underlying altered developmental trajectories and subsequent problematic alcohol use (Bogdan et al., 2023; Chassin et al., 2013).

### 4.6 Limitations in the current literature

The review highlights several key limitations in research on youth alcohol use and neurodevelopment. First, the measurement of alcohol use (i.e., frequency, quantity, and types of beverages) and definition of the severity of alcohol-use is inconsistent. The heterogeneity in metrics of alcohol use and related problems hinders the direct comparison of study findings and, therefore, limits a detailed understanding of how specific levels of alcohol use might be most impactful on youth brain development. For instance, the definitions of recent alcohol use across studies is varied, with some studies defining this as 3 months (Jones et al., 2023; Squeglia et al., 2014; Squeglia et al., 2015), whilst others define it as 1 month (El Marroun et al., 2021; Jones et al., 2023). Additionally, the literature often lacks explicit reporting on cannabis co-use, despite its high comorbidity with alcohol use among youth, making it difficult to assess its influence on outcomes (Subbaraman & Kerr, 2015). Furthermore, while two-timepoint datasets are considered longitudinal, they are limited in their ability to examine nonlinear trajectories and individual differences (Singer & Willett, 2003). Finally, large cohort studies like NCANDA deliberately oversampled high-risk youth (approximately 50% of participants), which limits the generalizability of the findings to the broader youth population.

### 4.7 Limitations of the review

This review has several limitations. First, it did not include grey literature (e.g., conference abstracts, posters, dissertations), which may lead to publication bias, potentially resulting in an overrepresentation of positive findings. Additionally, we did not conduct a quantitative meta-analysis to statistically measure the strength of effects across the literature due to significant inter-study heterogeneity in statistical methods and brain atlases used. Furthermore, the variability in reported metrics and the lack of standardised effect size reporting may limit the comparability of findings. As such, the broader impact of the findings of this review on clinical or policy applications is unclear.

## 5. Future directions

Emerging evidence underscores critical gaps in understanding the relationship between youth alcohol use and neurodevelopmental trajectories. Future longitudinal studies should prioritise methodological standardisation, particularly in defining alcohol consumption metrics (e.g., frequency, quantity), to facilitate robust cross-cohort comparisons. Furthermore, expanding research to include diverse, non-Western populations is also essential to establish the generalisability of findings.

Future studies should also aim to clarify the temporal dynamics of alcohol-related neurostructural changes, specifically, whether these alterations are transient or enduring. Further investigation is needed to establish how such changes associate with cognitive and functional outcomes (Cservenka & Brumback, 2017; Lees et al., 2020; Mota et al., 2013). Dose-response relationships require further exploration, particularly how varying levels of alcohol exposure correlate with structural alterations (e.g., frontal lobe grey matter reductions). Such research could inform evidence-based thresholds for safer consumption, especially when contextualised with environmental vulnerabilities such as familial history of AUD or childhood adversities. Finally, prospective studies assessing brain structure before and after alcohol initiation are critical to disentangle pre-existing vulnerabilities from alcohol-associated changes. Addressing these areas may contribute to the refinement of early intervention strategies and inform policy approaches aimed at reducing alcohol-related neurodevelopmental risks.

## 6. Conclusion

This systematic review synthesised findings from longitudinal studies examining the association between youth alcohol use and structural brain development. While the current evidence base is limited by both the number of studies and methodological heterogeneity, emerging findings suggest associations between alcohol use and GMV declines, particularly in temporal regions, in addition to attenuated WMV increases. Although preliminary, evidence from a limited number of studies suggests a potential dose-dependent relationship, whereby heavier consumption patterns may be linked to more pronounced GMV reductions. However, the relevance of these structural alterations for cognitive functioning in youth remains uncertain. Future research should address several key methodological considerations, including standardisation of alcohol use measurement across studies. Additionally, systematic investigation of potential moderating factors, including age of onset, sex, childhood trauma, substance co-use, and family history of alcohol use disorders, would help clarify individual differences in vulnerability. Addressing these methodological limitations, alongside more diverse longitudinal studies, will be critical for drawing more definitive conclusions about the impact of alcohol use on youth brain development.

## Data Availability

All data produced in the present work are contained in the manuscript

## Corresponding author

Prof Sarah Whittle, PhD

Orygen, and the Centre for Youth Mental Health, The University of Melbourne Address: Orygen, 35 Poplar Avenue, Parkville VIC 3052, Australia

Declaration of Generative AI and AI-assisted technologies in the writing process

No generative AI or AI-assisted technologies were used during the preparation of this work.

