## Supplementary Material for "The impact of alcohol use in youth neurodevelopment: A systematic review of longitudinal structural neuroimaging studies"

**1.1 Additional data handling**

Additional data handling was required for several reasons. Firstly, to address the variability in how findings were reported, when studies examined multiple effects for each imaging metric, specifically group and group-by-time, each effect was reported independently. Secondly, El Marroun et al. (2020) and Jones et al. (2023) present findings from separate cohorts; the findings from each cohort are reported individually and alphabetically distinguished (e.g., El Marroun et al 2021 (a) would refer to findings from the BrainScale cohort). Refer to Table 1 to see the alphabetical representation of each cohort in these two studies. Lastly, Hua et al. (2022) met our criteria for examining alcohol effects over multiple time points, but with only a 5-week interval between them. The results from this study were considered, noting the unusually short duration between assessments, which may not fully capture the effects on neurodevelopment (Squeglia et al., 2009).

**1.2 PRISMA Flowchart**

The PRISMA flowchart in Figure 1 describes the overall screening process using the systematic review tool, Covidence (www.covidence.org). Keywords used for the search included terms related to alcohol use behaviours (e.g., "alcohol use," "underage drinking," "binge drinking"), youth population (e.g., "adolescent," "teen," "young adult"), neuroimaging and brain development measures (e.g., "brain development," "gray matter," "white matter," "cortical thickness," "MRI"), and longitudinal study designs (e.g., "longitudinal studies," "follow-up studies," "prospective studies"). The searches retrieved 854 studies, of which 11 duplicates were removed. Then, the titles and abstracts of 843 studies were independently screened by two researchers (D.D.R and E.Mc) as per the inclusion and exclusion criteria, which led to the exclusion of 814 articles. A full-text review of 29 studies led to the omission of 15 studies due to meeting the exclusion criteria. After full text review, 14 articles were retained as meeting inclusion criteria for the systematic review. One additional study was then included after cross-referencing the selected articles.

**Identification**

References from other sources **(n = 2)**

Citation searching (n =2 )

Studies screened **(n = 845)**

Studies sought for retrieval **(n = 32)**

Studies assessed for eligibility **(n = 29)**

References removed **(n = 11)**

Duplicates identified manually (n = 0)

Duplicates identified by Covidence (n = 11)

Marked as ineligible by automation tools (n = 0)

Studies excluded **(n = 813)**

Studies excluded **(n = 13)**

Wrong study design (n = 5)

Not empirical study (n = 6)

Wrong patient population (n = 2)

Studies included in review **(n = 16)**

**Screening**

Studies from databases/registers **(n = 856)**

**Included**

**Figure S1:** PRISMA Flowchart summarising the inclusion and exclusion of the studies reviewed in this systematic review

**1.3 MRI data acquisition and analysis parameters**

The reviewed articles used a variety of analysis approaches. Ten studies used the automated segmentation FreeSurfer V 6.0 and segmented 170 regions and sub-regions (Jones et al., 2023; Sun et al., 2023 Infante et al., 2022; Luo et al., 2022; Philips et al., 2021; Hua et al., 2020; Pfefferbaum et al., 2018; Infante et al., 2018; Jacobus et al., 2016; Squeglia et al., 2014; Luciana et al., 2013). Three studies included a whole brain analysis using FreeSurfer (Sun et al., 2023; Perez-Garcia et al., 2022; Luciana et al., 2013); while one study used ROBEX (Squeglia et al., 2014). One study examined specifically cerebellum total and subregional volumes of the software used SUIT (Sullivan et al., 2020). Finally, one study examined whole brain volume and GMV using voxel-based morphometry (Meda et al., 2017).

***Acquisition hardware***

Several MRI parameters were used across the reviewed studies. MRI scanners used varied across these studies, and it was most consistently the GE Discovery MR 750 scanner (Sun et al., 2023; Infante et al., 2022, Luo et al., 2022; Phillips et al., 2021; Sullivan et al., 2020; Pfefferbaum et al 2018; Marroun et al., 2021); and Siemens TIM TRIO scanner in six studies (Sun et al., 2023; Infante et al., 2022; Luo et al., 2022; Phillips et al., 2022; Sullivan et al., 2020; Pfefferbaum et al., 2018). Of these, the six studies that used data from the NACANDA consortium used both the GE Discovery MR 750 and the Siemens TIM TRIO scanners, with their data collected across 5 sites (Infante et al., 2022; Jones et al., 2023; Luo et al., 2022; Pfefferbaum et al., 2018; Phillips et al., 2021; Sullivan et al., 2020; Sun et al., 2023). The Philips Achieva Scanner is used in two studies (Perez-Garcia et al., 2022; Marroun et al., 2021) and the 3-Tesla CXK4 short bore Excite-2 scanner was used in two studies (Squeglia et al., 2015; Squeglia et al., 2014). The Siemens Allegra 3T scanner is used in one study (Meda et al., 2017). Finally, the Siemens Trio 3T scanner features in one study (Luciana e al., 2013)

***MRI acquisition metrics***

The most used scanner across the 16 studies was the 3 Tesla General Electric (GE) Discovery MR750 and Siemens TIM TRIO (n=8). The typical parameters for these scanners include a field of view (FOV) of 240 mm × 240 mm, acquisition matrix of 256 × 256, and slice thickness of 1.2 mm. The GE scanners often used an IR-FSPGR sequence with TR/TI/TE parameters of 5.912/400/1.932 ms, a flip angle of 11°, and 146 slices. Meanwhile, the Siemens scanners used an MPRAGE sequence with TR/TI/TE parameters of 1900/900/2.92 ms, a flip angle of 9°, and 160 slices. Studies by Hua et al. (2020), Pfefferbaum et al. (2018), and Luciana et al. (2013) did not report key MRI parameters such as TE, TI, voxel size, matrix size, or slice number. Information about the MRI acquisition parameters for all studies can be found in Supplementary Table S1.

| **Table S1: Overview of MRI data acquisition parameters across the reviewed studies** | | | | | | | | | | | | | | | | | |
| --- | --- | --- | --- | --- | --- | --- | --- | --- | --- | --- | --- | --- | --- | --- | --- | --- | --- |
| **Study** | **Country of Origin** | **Type of analysis** | **Scanner Model** | ***TE, ms*** | ***TI, ms*** | ***TR, ms*** | ***FOV mm3*** | ***Flip angle size (degree)*** | **Voxel size, mm** | **Matrix size** | **Sequece type** | **NEX** | **plane** | **head coil** | **SLICE N** | **Slick (mm)** | **Software pipeline** |
| **Jones et al., 2023 (a)** | USA | ROI | GE MR750 (3T) | 1.932 | 400 | 5.912 | 240 x 240 | 11 | _ | 256 x 256 | IR-FSPGR | _ | Sagittal | 8- channel | 146 | _ | FreeSurfer 6.0, longitudinal segmentation |
|  |  |  | Siemens TIM TRIO | 2.92 | 900 | 1900 | 240 x 240 | 9 | _ | 256 x 256 | MPRAGE | _ | Sagittal | 8- channel | 160 | _ |  |
| Jones et al., 2023 (b) | USA | ROI | Siemens TIM TRIO | 3.58–3.61 | 900 | 2300 | _ | 10 | 1.1 × 1 × 1 | _ | _ | _ | Sagittal | 12-channel | 160 | _ | FreeSurfer 6.0, longitudinal segmentation |
| Sun et al., 2023 | USA | Whole brain voxelwise | GE MR750 +D2:R3(3T) | 1.932 | 400 | 5.912 | 240 x 240 | 11 | _ | 256 x 256 | IR-FSPGR | _ | Sagittal | 8- channel | 146 | _ | FreeSurfer v6.0 longitudinal stream, ComBat |
|  |  |  | Siemens TIM TRIO | 2.92 | 900 | 1900 | 240 x 240 | 9 | _ | 256 x 256 | MPRAGE | _ | Sagittal | 8- channel | 160 | _ |  |
| Perez Garcia et al., 2022 | Spain | Whole brain voxelwise & ROI | 3T Achieva Philips | 3.4 | - | 7.7 | 240 | 8 | 0.8 | - | 3D turbo field-echo | _ | Transverse | 32-channel SENSE head coil | 200 | _ | FreeSurfer 6.0, cross-sectional and longitudinal pipelines |
| [Infante et al., 2022](https://drive.google.com/file/d/1-Q0Z0RfvWuR0ei8wWXwg66t3bAt3e5wK/view?usp=sharing) | USA | ROI | GE MR750 (3T) | 1.932 | 400 | 5.912 | 240 x 240 | 11 | _ | 256 x 256 | IR-FSPGR | _ | Sagittal | 8- channel | 146 | _ | FreeSurfer cross-sectional and longitudinal streams |
|  |  |  | Siemens TIM TRIO | 2.92 | 900 | 1900 | 240 x 240 | 9 | _ | 256 x 256 | MPRAGE | _ | Sagittal | 8- channel | 160 | _ |  |
| Luo et al., 2022 | USA | ROI | GE MR750 (3T) | 1.932 | 400 | 5.912 | 240 x 240 | 11 | _ | 256 x 256 | IR-FSPGR | _ | Sagittal | 8- channel | 146 | _ | FreeSurfer v6.0 cross-sectional and longitudinal streams |
|  |  |  | Siemens TIM TRIO | 2.92 | 900 | 1900 | 240 x 240 | 9 | _ | 256 x 256 | MPRAGE | _ | Sagittal | 8- channel | 160 | _ |  |
| Phillips et al., 2021 | USA | ROI | GE MR750 (3T) | 1.932 | 400 | 5.912 | 240 x 240 | 11 | 1.2 x 0.9375 | 256 x 256 | IR-FSPGR | 1 | _ | 8- channel | 146 | _ | FreeSurfer v6.0, longitudinal segmentation pipeline |
|  |  |  | Siemens TIM TRIO | 2.92 | 900 | 1900 | 240 x 240 | 9 | 1.2 x 0.9375 | 256 x 256 | MPRAGE | 1 | _ | 12- channel | 160 | _ |  |
| Sullivan et al., 2019 | USA | ROI | GE MR750 (3T) | 1.932 | 400 | 5.912 | 240 x 240 | 11 | 1.2 x 0.9375 | 256 x 256 | IR-FSPGR | 1 | _ | 8- channel | 146 | _ | FreeSurfer v6.0, longitudinal segmentation pipeline |
|  |  |  | Siemens TIM TRIO | 2.92 | 900 | 1900 | 240 x 240 | 9 | 1.2 x 0.9375 | 256 x 256 | MPRAGE | 1 | _ | 12- channel | 160 | _ |  |
| Hua et al., 2021 | USA | ROI | Siemens Trio 3T | 2.92 | 900 | 1900 | 256 x 256 | 9 | _ | 256 x 256 | IMPRAGE | _ | _ | 8 | _ | 1 | FreeSurfer v6.0 (segmentation unknown) |
| [EL Marroun et al., 2021 (a)](https://drive.google.com/file/d/1EBAt1PqbF8Ih5GBS4rXoRvY1oErvdRdE/view?usp=sharing) | Netherlands | ROI | Philips Achieva Scanner (1.5 T) | 4.6 | - | 30 | - | 30 | 1 × 1 × 1.2 | 256 × 256 | 3D | _ | Coronal | _ | 160 | 1.2 | FreeSurfer v6.0 for cross-sectional and longitudinal analysis |
| EL Marroun et al., 2021 (b) | Netherlands | ROI | Philips Achieva TX 3.0T scanner (3T) | 4.59 | - | 9.76 | 224 × 177 | 8 | 0.875 × 0.875 × 1.2 | - | 3D | _ | _ | _ | 140 | _ | FreeSurfer v6.0 for cross-sectional and longitudinal analysis |
| EL Marroun et al., 2021 (c) | Netherlands | ROI | GE MR750 (3T) | 3.4 | 600 | 8.77 | 220 × 220 | 10 | 1 × 1 × 1 | 220 × 220 | IR-prepared | - | - | 8-channel | 230 | 1 | FreeSurfer v6.0 for cross-sectional and longitudinal analysis |
| Pfefferbaum et al., 2018 | USA | ROI | GE MR750 (3T) | 1.932 | 400 | 5.912 | 240 x 240 | 11 | _ | 256 x 256 | IR-FSPGR | _ | Sagittal | 8- channel | 146 | _ | FreeSurfer cross-sectional and longitudinal streams |
|  |  |  | Siemens TIM TRIO | 2.92 | 900 | 1900 | 240 x 240 | 9 | _ | 256 x 256 | MPRAGE | _ | Sagittal | 8- channel | 160 | _ |  |
| Infante et al., 2018 | USA | ROI | GE 3.0 Tesla CXK4 Excite-2 | _ | _ | _ | _ | _ | _ | _ | _ | _ | _ | 8 -channel | _ | _ | FreeSurfer v5.1 longitudinal stream |
| Jacobus et al., 2016 | USA | ROI | GE 3.0 Tesla CXK4 Excite-2 | _ | _ | _ | ` | 12 | 0.94 x 0.94 x 1 | 256 x 256 | _ | _ | _ | 8 -channel | 176 | _ | Freesurfer v5.1 |
| Meda et al., 2017 | USA | Whole brain voxelwise | Siemens Allegra 3T | 2.74 | 900 | 2300 | 176 x 256 | 8 | 1 | 176 x 256 | MPRAGE | _ | Sagittal | _ | 176 | _ | Symmetric diffeomorphic modelling in SPM12 |
| Squeglia et al., 2015 | USA | ROI | 3-Tesla CXK4 short-bore Excite-2 | 4.8 | - | 20 | 24 | 12 | 0.943 x 0.94 | 256 x 256 | 3D T1-weighted anatomical MRI | - | Sagittal | 8-channel | 176 | _ | FSL's FAST tool, SRI24 parcellation maps |
| Squeglia et al., 2014 | USA | ROI | 3-Tesla CXK4 short bore Excite-2 | 4.8 | - | 20 | 24 | 12 | 0.943 x 0.94 | 256 × 256 | 3D T1-weighted anatomical MRI | - | Sagittal | 8-channel | 176 | _ | QUARC, FreeSurfer 4.5.0 |
| Luciana et al., 2013 | USA | ROI | Siemens 3 T scanner | 3.65 | 1100 | 2530 | 256 | 7 | 1.0 x 1.0 | 256 x 256 | IMPRAGE | _ | coronal | 8-channel | 240 | _ | FreeSurfer software suite v4.5.0, QUARC |
| Missing data is marked with ‘_’ | | | | | | | | | | | | | | | | | |

**1.4 Data synthesis**

Due to study heterogeneity, we employed a label-based meta-analysis approach, counting how many times particular brain regions were detected across studies. Author-defined brain regions were grouped into standard lobes (e.g., inferior frontal gyrus → frontal lobe) and subcortical regions (see Table S2 for examples). Consistency was defined as >50% of studies reporting significant findings in a particular region, expressed as percentages with fractional equivalents (e.g., "frontal lobe (60%, 3/5)"). Results were reported by cohort rather than by study, treating each cohort as an independent finding.

| **Table S2: Brain region categorisation for studies** | | | | | | | | | | | | | | |
| --- | --- | --- | --- | --- | --- | --- | --- | --- | --- | --- | --- | --- | --- | --- |
| ROI name | Perez Garcia | Luo | Sullivan | Hua | El Marroun A | El Marroun B | El Marroun C | Pfefferbaum | Meda | Squeglia 2015 | Squeglia 2014 | **Total Sig** | **Total non-sig** | **Total mentions** |
| **Parietal lobe** | _ | 1 | _ | _ | _ | _ | _ | 0 | _ | 0 | _ | 1 | 2 | 3 |
| angular | _ | _ | _ | _ | _ | _ | _ | _ | _ | 0 | _ | 0 | 1 | 1 |
| supramarginal | _ | 1 | _ | _ | _ | _ | _ | 0 | _ | 0 | 0 | 1 | 3 | 4 |
| superior | _ | 0 | _ | _ | _ | _ | _ | 0 | _ | 0 | 0 | 0 | 4 | 4 |
| post central | _ | _ | _ | _ | _ | _ | _ | _ | _ | 0 | _ | 0 | 1 | 1 |
| precuneus | _ | 0 | _ | _ | _ | _ | _ | 0 | _ | 0 | 0 | 0 | 4 | 4 |
| **Frontal lobe** | 0 | 0 | _ | _ | 1 | 1 | 0 | 1 | 1 | 1 | 0 | 5 | 4 | 9 |
| inferior frontal gyrus | 0 | _ | _ | _ | _ | _ | _ |  | _ | _ | _ | 0 | 1 | 1 |
| superior frontal gyrus | 0 | 0 | _ | _ | 1 | 0 | 0 | 1 | _ | _ | 0 | 2 | 5 | 7 |
| lateral frontal cortex | _ | _ | _ | _ | 1 | 0 | 0 | 0 | _ | 1 | _ | 2 | 3 | 5 |
| medial frontal cortex | _ | _ | _ | _ | _ | _ | _ | 0 | _ | 1 | _ | 1 | 1 | 2 |
| frontopolar | 0 | 0 | _ | _ | _ | _ | _ | 0 | 0 | _ | 0 | 0 | 5 | 5 |
| caudal middle frontal gyrus | _ | 0 | _ | _ | 1 | 0 | 0 | 1 | _ | _ | 0 | 2 | 4 | 6 |
| rostral middle frontal gyrus | 0 | 0 | _ | _ | 1 | 1 | 0 | 0 | 0 | _ | 0 | 2 | 6 | 8 |
| precentral | _ | 0 | _ | _ | _ | _ | _ | 0 | 0 | _ | 0 | 0 | 4 | 4 |
| paracentral | _ | 0 | _ | _ | _ | _ | _ | 0 | 1 | _ | 0 | 1 | 3 | 4 |
| OFC medial | _ | 0 | _ | _ | 0 | 1 | 0 | 0 | _ | _ | 0 | 1 | 5 | 6 |
| OFC lateral | _ | 0 | _ | _ | _ | _ | _ | 0 | _ | _ | 0 | 0 | 3 | 3 |
| IFG pars opercularis | _ | 0 | _ | _ | _ | _ | _ | 0 | 1 | _ | 0 | 1 | 3 | 4 |
| IFG orbitalis | _ | 0 | _ | _ | _ | _ | _ | 0 | 1 | _ | 0 | 1 | 3 | 4 |
| IFG trangularis pars | _ | 0 | _ | _ | _ | _ | _ | 0 | 1 | _ | 0 | 1 | 3 | 4 |
| **Temporal lobe** | _ | 1 | _ | _ | _ | _ | _ | 0 | 0 | 1 | 1 | 3 | 2 | 5 |
| superior | _ | 1 | _ | _ | _ | _ | _ | 0 | 0 | 1 | 0 | 2 | 3 | 5 |
| middle | _ | 0 | _ | _ | _ | _ | _ | 0 | 0 | 1 | 1 | 2 | 3 | 5 |
| inferior | _ | 0 | _ | _ | _ | _ | _ | 0 | 0 | 1 | 1 | 2 | 3 | 5 |
| pole | _ | 0 | _ | _ | _ | _ | _ | 0 | 0 | 1 | 0 | 1 | 4 | 5 |
| banks | _ | 0 | _ | _ | _ | _ | _ | 0 | 0 | 1 | 0 | 1 | 4 | 5 |
| entorhinal | _ | 0 | _ | _ | _ | _ | _ | 0 | 0 | 1 | 0 | 1 | 4 | 5 |
| transverse | _ | 0 | _ | _ | _ | _ | _ | 0 | 0 | 1 | 0 | 1 | 4 | 5 |
| **Cingulate** | _ | _ | _ | _ | 0 | 0 | 0 | 1 | 1 | 0 | 0 | 2 | 5 | 7 |
| caudal anterior | _ | _ | _ | _ | 0 | 0 | 0 | 0 | 0 | 0 | 0 | 0 | 7 | 7 |
| rostral anterior | _ | _ | _ | _ | 0 | 0 | 0 | 0 | 0 | 0 | 0 | 0 | 7 | 7 |
| posterior | _ | _ | _ | _ | 0 | 0 | 0 | 1 | _ | 0 | 0 | 1 | 6 | 7 |
| middle | _ | _ | _ | _ | _ | _ | _ | _ | _ | 0 | 0 | 0 | 2 | 2 |
| isthmus | 0 | _ | _ | _ | _ | _ | _ | 0 | 1 | 0 | 0 | 1 | 4 | 5 |
| anterior | _ | 0 | _ | _ | _ | _ | _ | 0 | 1 | 0 | 0 | 1 | 4 | 5 |
| **Insula** | _ | 0 | _ | _ | _ | _ | _ | 0 | _ | 0 | 0 | 0 | 4 | 4 |
| **Occipital lobe** | _ | 0 | _ | _ | _ | _ | _ | 0 | _ | 0 | 0 | 0 | 4 | 4 |
| lingual | _ | 0 | _ | _ | _ | _ | _ | 0 | _ | 0 | 0 | 0 | 4 | 4 |
| lateral | _ | 0 | _ | _ | _ | _ | _ | 0 | _ | 0 | 0 | 0 | 4 | 4 |
| pericalcarine | _ | 0 | _ | _ | _ | _ | _ | 0 | _ | 0 | 0 | 0 | 4 | 4 |
| fusiform | _ | 0 | _ | _ | _ | _ | _ | 0 | _ | 0 | 0 | 0 | 4 | 4 |
| cuneus | _ | 0 | _ | 0 | 1 | 0 | 0 | 0 | 1 | _ | 0 | 2 | 6 | 8 |
| **Hippocampus** | _ | 0 | _ | 0 | 1 | 0 | 0 | 0 | 0 | _ | 0 | 1 | 7 | 8 |
| prosubiculum | _ | 0 | _ | 0 | 1 | 0 | 0 | 0 | 0 | _ | 0 | 1 | 7 | 8 |
| subiculum proper | _ | 0 | _ | 0 | 1 | 0 | 0 | 0 | 0 | _ | 0 | 1 | 7 | 8 |
| presubiculum | _ | 0 | _ | 0 | 1 | 0 | 0 | 0 | 1 | _ | 0 | 2 | 6 | 8 |
| parasubiculum | _ | _ | 1 | _ | _ | _ | _ | _ | _ | _ | 0 | 1 | 1 | 2 |
| **Cerebellum** | _ | _ | 1 | _ | _ | _ | _ | _ | _ | _ | 0 | 1 | 1 | 2 |
| I | _ | _ | 1 | _ | _ | _ | _ | _ | _ | _ | 0 | 1 | 1 | 2 |
| IV | _ | _ | 1 | _ | _ | _ | _ | _ | _ | _ | 0 | 1 | 1 | 2 |
| V | _ | _ | 1 | _ | _ | _ | _ | _ | _ | _ | 0 | 1 | 1 | 2 |
| VI | _ | _ | 1 | _ | _ | _ | _ | _ | _ | _ | 0 | 1 | 1 | 2 |
| Crus I | _ | _ | 1 | _ | _ | _ | _ | _ | _ | _ | 0 | 1 | 1 | 2 |
| Crux lI | _ | _ | 1 | _ | _ | _ | _ | _ | _ | _ | 0 | 1 | 1 | 2 |
| VIIb | _ | _ | 1 | _ | _ | _ | _ | _ | _ | _ | 0 | 1 | 1 | 2 |
| Vermis | _ |  | _ | _ | 1 | 0 | 0 | _ | _ | _ | _ | 1 | 2 | 3 |
| **Amygdala** | 0 | 0 | _ | _ | 0 | 1 | 0 | _ | 0 | _ | 1 | 2 | 5 | 7 |
| **Striatum** | 0 | 0 | _ | _ | 0 | 1 | 0 | _ | 0 | _ | 0 | 1 | 6 | 7 |
| Accumbens | 0 | 0 | _ | _ | 0 | 0 | 0 | _ | 0 | _ | 1 | 1 | 6 | 7 |
| caudate | 0 | 0 | _ | _ | 0 | 0 | 0 | _ | 0 | _ | 0 | 0 | 7 | 7 |

**2. Results**

**2.1 Main effect of alcohol use**

As shown in Table S3, the main effect of alcohol use was reported for nine cohorts (El Marroun et al., 2021; Hua et al., 2020; Infante et al., 2018; Jones et al., 2023; Luo et al., 2022; Sullivan et al., 2020). The most consistently studied imaging metric was GMV (n=8), followed by WMV (n=3) and by cortical thickness (n=1).

***GMV:*** GMV differences between alcohol users and controls were reported for eight cohorts. The most consistently reported significant findings were observed within the frontal lobe (60%, 3/5) and the superior temporal cortex (100%, 3/3), where smaller GMV was reported in alcohol users compared to non-users/low level users.

***WMV:*** WMV differences between alcohol users and controls were investigated in three cohorts. El Marroun et al. (2021) found a significant reduction in WMV in the central white matter between alcohol users and non-users in cohort A, and no group differences in cohorts B and C.

***Cortical thickness:*** Only Sun et al. (2023) investigated differences in thickness between alcohol users and controls. The study reported that alcohol use compared low use was associated with smaller cortical thickness in the superior and middle frontal regions of the brain, but not in the other ROI investigated.

| **Table S3: Main effect of group (alcohol users vs controls)** | | | | | |
| --- | --- | --- | --- | --- | --- |
| **Studies** | **Type of analysis** | **Groups compared** | **Neurometric** | **significant** | **non-significant** |
| Jones et al., 2023 (a) | ROI | alcohol vs HC | GMV | *SMALLER* in alcohol vs HC in hippocampus (whole) | **amygdala, striatum** (accumbens/caudate/putamen), **thalamus, pallidum** |
| Jones et al., 2023 (b) |  | alcohol vs HC | GMV | *SMALLER* in alcohol vs HC in hippocampus (whole) | **amygdala, striatum** (accumbens/caudate/putamen), **thalamus, pallidum** |
| Sun et al., 2023 | Whole brain | Heavy vs low | thickness | *Smaller in heavy use vs HC*  in **frontal** (superior, medial), **temporal** (superior, middle, inferior), **cingulate** (anterior, middle, posterior), **insula**, **occipital** (lingual, lateral, pericalcarine, fusiform), **parietal** (cuneus, angular, supramarginal) | n/a |
| Luo et al., 2022 | ROI | alcohol vs HC | GMV | *SMALLER in alcohol vs HC* in **frontal** (caudal), **temporal** (superior), **occipital** (fusiform), **parietal** (supramarginal) | **frontal** (OFC, pars opercularis/orbitalis/triangularis pole, superior, caudal/rostral middle, pre/para/post-central), **hippocampus** (presubiculum, subiculum proper, presubiculum, parasubiculum), **temporal** (pole, banks, entorhinal, middle/inferior, transverse), **cingulate** (caudal/rostral/posterior/anterior), **insula**, **occipital** (pericalcarine, cuneus, lateral, lingual), **striatum** (accumbens/caudate/putamen), **parietal** (superior, precuneus) |
| Sullivan et al., 2020 | ROI | alcohol vs HC | GMV | *SMALLER in alcohol vs HC* in **cerebellum** lobes (I-IV, V, VI, Crux I/II, VIIb) & vermis | _ |
| El Marroun et al., 2020 (a) | Whole brain and ROI | alcohol vs HC | GMV | *SMALLER in alcohol vs HC*  in **frontal** (superior), **hippocampus**, **amygdala** | **frontal** (OFC, lateral, caudal middle, rostral middle), **cingulate** (caudal/rostral/posterior, isthmus), **striatum** (accumbens/caudate/putamen) |
|  |  |  | WMV | *SMALLER increase in alcohol vs HC*  in **global white matter** | **_** |
| El Marroun et al., 2020 (b) |  | alcohol vs HC | GMV | *SMALLER in alcohol vs HC*  in **frontal** (superior, rostral middle, medial orbital), **cingulate** (posterior, isthmus), **striatum** (accumbens) | **frontal** (OFC, lateral, caudal), **hippocampus**, **cingulate** (caudal/rostral), **amygdala**, **striatum** (caudate/putamen) |
|  |  |  | WMV | **_** | **global white matter** |
| El Marroun et al., 2020 (c) |  | alcohol vs HC | GMV | **_** | **frontal** (OFC, lateral, superior, caudal/rostral middle), **hippocampus**, **cingulate** (caudal/rostral/posterior/isthmus), **amygdala**, **striatum** (accumbens/caudate/putamen) |
|  |  |  | WMV | **_** | **global white matter** |
| Infante et al., 2018 | ROI | binge drinking vs non-binge | GMV | *SMALLER in alcohol vs HC*  in **temporal** (superior, inferior), **occipital** (fusiform) | **frontal** (OFC, pars opercularis/orbitalis/triangularis pole, medial, caudal middle, rostral middle, pre/para/post-central), **temporal** (pole, banks, entorhinal, middle, transverse), **cingulate** (caudal/rostral/anterior/ posterior), **insula**, **occipital** (pericalcarine, cuneus, lateral, lingual), **parietal** (supramarginal, superior, precuneus) |
| **Abbreviations:** WMV = white matter volume, GMV= grey matter volume, ‘_’ = none, HC = healthy controls, OFC= orbitofrontal cortex, IFG= inferior frontal gyrus; n/a = Not applicable as the analysis was whole brain | | | | | |

**2.3. Additional Moderators**

Eight studies investigated additional variables as moderators of the longitudinal relationship between alcohol use and structural neurodevelopment in youth (Infante et al., 2018; Infante et al., 2022; Jacobus et al., 2016; Jones et al., 2023; Luo et al., 2022; Pfefferbaum et al., 2018; Sullivan et al., 2020; Sun et al., 2023). These variables include co-use of alcohol and cannabis (n=3), sex (n=2), trauma (n=2), and family history of alcohol use or alcohol use disorder (AUD) (n=3). The main effects of these moderating variables are not described here as the focus is on the interactive effects with time, and summarising the main effects is beyond the scope of this review.

***Alcohol and cannabis co-use***

The longitudinal effects of alcohol and cannabis co-use were examined in three cohort studies (Infante et al., 2018; Jacobus et al., 2016; Luo et al., 2022). Infante et al. (2018) and Jacobus et al. (2016) found that alcohol-only users showed greater reductions in the surface area and cortical thickness of the frontal and parietal lobes over time compared to co-users. Conversely, Luo et al. (2022) reported that co-users experienced significantly greater declines in brain volume in the frontal, temporal, and parietal lobes compared to both non-users and alcohol-only users.

***Sex***

Two cohorts investigated sex-by-alcohol-by-time interactions (Infante et al., 2022; Jones et al., 2023). Jones et al. (2023) found significant interactions by sex, whereby alcohol use was significantly associated with reduced volume in the caudate and thalamus in females over time. In males, there was no association between alcohol use and volume in any of these regions. On the other hand, Infante et al. (2022) found no significant interactions between binge drinking, sex and time.

***Trauma***

Two cohorts reported on the interaction between alcohol use, trauma and time on brain structure (Infante et al., 2022; Sun et al., 2023). Both studies reported non-significant interactions in all ROI for cortical thickness (Sun et al., 2023) and GMV (Infante et al., 2022).

***Familial history of alcohol use***

The interaction between family history of alcohol use, youth alcohol use and time on brain structure was examined in three cohort studies (Pfefferbaum et al., 2018; Sullivan et al., 2020; Sun et al., 2023). Firstly, Pfefferbaum et al. (2018) found that heavy drinkers with a positive family history of alcoholism showed steeper declines in parietal, occipital, and total grey matter volumes compared to those with a negative family history. Similarly, Sullivan et al. (2020) observed that among alcohol users, those with a positive family history showed a steeper decline in the GMV of cerebellar lobules I to IV compared to those with a negative family history. Conversely, Sun et al. (2023) found no significant interactions for any brain regions.
